# COVID-19 Risk Factors and Mortality among Native Americans

**DOI:** 10.1101/2021.03.13.21253515

**Authors:** Katherine Leggat-Barr, Fumiya Uchikoshi, Noreen Goldman

## Abstract

**BACKGROUND:** Academic research on the disproportionate impact of COVID-19 among Native Americans has largely been restricted to particular indigenous groups or reservations.

**OBJECTIVE:** We estimate COVID-19 mortality for Native Americans relative to other racial/ethnic groups and explore how state-level mortality is associated with known risk factors. METHODS: We use the Standard Mortality Ratio (SMR), adjusted for age, to estimate COVID-19 mortality by racial/ethnic groups for the U.S. and 16 selected states that account for three-quarters of the Native American population. The prevalence of risk factors is derived from the American Community Survey and the Behavioral Risk Factor Surveillance System.

**RESULTS:** The SMR for Native Americans greatly exceeds those for Black and Latino populations and varies enormously across states. There is a strong positive correlation across states between the share of Native Americans living on a reservation and the SMR. The SMR for Native Americans is highly correlated with the income-poverty ratio, the prevalence of multigenerational families, and health insurance (excluding the Indian Health Service). Risk factors associated with socioeconomic status and co-morbidities are generally more prevalent for Native Americans living on homelands, a proxy for reservation status, than for those living elsewhere.

**CONCLUSIONS:** Most risk factors for COVID-19 are disproportionately high among Native Americans. Reservation life appears to increase the risk of COVID-19 mortality.

**CONTRIBUTION:** We assemble and analyze a broader set of COVID-19-related risk factors for Native Americans than previous studies, a critical step toward understanding the exceptionally high COVID-19 death rates in this population.

## 1. Introduction

One of the many egregious consequences of the COVID-19 pandemic has been the disproportionate impact of morbidity and mortality on already disadvantaged populations (Thakur et al. 2020; Andrasfay and Goldman 2021). Although the full effect of the coronavirus for all racial and ethnic groups has yet to be determined, high quality estimates for Native Americans remain particularly scarce. Despite extensive media attention to Native Americans, academic research on COVID-19 in this population has been much less common than for Black and Latino Americans, and along with media reports, has been largely restricted to particular indigenous groups or reservations. This limitation is due in large part to the deficiencies and paucity of data for Native Americans (Hatcher et al. 2020).

In U.S. Census data, Native Americans are individuals who self-identify as American Indian or Alaska Native (AI/AN) alone or in conjunction with other racial or ethnic groups. They comprise about 6 million individuals or two percent of the U.S. population, with about half of this group identifying as single-race Native American (Norris, Vines, and Hoeffel 2012). The vast majority of Native Americans self-identify as American Indian rather than Alaska Native.^1^ We use the term Native American to refer to estimates for the combined group AI/AN.

As with other marginalized groups, high rates of vulnerability to COVID-19 among Native Americans have been attributed to long-standing structural inequalities and racism that include poverty, inadequate health care and health insurance, a high prevalence of co-morbidities that increase susceptibility to COVID-19, and poor living conditions including inadequate plumbing and crowded living quarters (Kakol, Upson, and Sood 2020). Some or all of these factors may be especially prevalent on Native American reservations (Dewees and Marks, 2017; Yellow Horse, Deschine Parkhurst, and Huyser 2020; Yellow Horse, Yang, and Huyser 2021). Nevertheless, the importance of these risk factors for COVID-19 mortality among Native Americans remains largely speculative.

Our paper has several objectives. The first is to provide improved estimates of COVID-19 death rates for Native Americans in comparison with the Black, Latino and White populations. The most frequently reported measures, such as the proportion of the group dying from COVID-19 in relation to the group’s representation in the population, fail to adjust for the large differences in underlying age distributions across racial and ethnic groups. There are, however, several recent estimates that account for these age differences (Bassett, Chen, and Krieger 2020; Xu et al. 2021).

The second goal is to determine the prevalence and potential importance of risk factors that may be related to COVID-19 mortality. We measure a broader set of risk factors compared with most earlier studies on Native American vulnerability: poverty, frequency of multigenerational households and residential crowding, health insurance coverage, frontline worker status, smoking status, the prevalence of seven health conditions that may increase the severity of COVID-19 infection and risk of death, and state policies to mitigate viral transmission. We estimate these COVID-19 death rates and prevalence of risk factors at both a national level and for the 16 states with the largest Native American populations: Alaska, Arizona, California, Colorado, Florida, Michigan, Minnesota, Montana, New Mexico, New York, North Carolina, Oklahoma, South Dakota, Texas, Washington and Wisconsin. Slightly more than three-quarters of Native Americans in the United States reside in these 16 states.

The final objective of this paper is to assess whether vulnerability to COVID-19 varies between Native Americans who live on vs. off reservations and whether reservation status is associated with COVID-19 death rates across states.

In order to address these questions, we analyze several data sources: provisional counts of COVID-19 deaths during 2020 from the Centers for Disease Control, recent population counts for the U.S., and potential risk factors from both the American Community Survey (ACS) and the Behavioral Risk Factor Surveillance Survey (BRFSS).

## 2. Background

SARS-CoV-2, the virus responsible for COVID-19, is transmitted primarily through respiratory droplets and aerosols when an infected person sneezes, coughs, talks or breathes in close proximity to others (Wilson, Corbett, and Tovey 2020). Research has identified a diverse set of socioeconomic and health-related risk factors that increase the likelihood of viral infection, illness severity, hospitalization and mortality. Many of these factors are likely to be especially prevalent among Native Americans, rendering them socially vulnerable to the coronavirus (Hathaway 2020; Yellow Horse, Yang, and Huyser 2021).

Both individual and community-level poverty are risk factors for COVID-19 transmission and severity (Patel et al. 2020; Williamson et al. 2020). Poverty status operates through multiple exposures including employment status and job characteristics, housing and living conditions, health-related behaviors and health status. Studies have pointed to likely higher risks among frontline and essential workers, who are often drawn disproportionately from disadvantaged racial and ethnic groups, and have jobs involving close indoor contact with others, often without adequate protective measures and without the possibility of being performed remotely (Goldman et al. 2021; Selden and Berdahl 2020). Frequent reliance on public transport exacerbates the infection risks faced by these workers (McLaren 2021). Because household transmission has been a significant source of COVID-19 spread (Haroon et al. 2020; Shen et al. 2020), multigenerational housholds, particularly those in crowded living spaces, enhance COVID-19 transmission and are particularly dangerous for the elderly (Jing et al. 2020; Chen and Kreiger 2021).

One reason for the much higher death rates among older individuals is the increasing prevalence with age of co-morbidities that are believed to raise the risk of dying from the virus. According to the Centers for Disease Control, the health-related conditions in the highest risk category include obesity, cardiovascular disease, cancer, chronic obstructive pulmonary disease, (type 2) diabetes, and chronic kidney disease (Centers for Disease Control and Prevention 2021b). Although there is some evidence that asthma is also a risk factor for COVID-19 severity and fatality, findings to date are mixed (Williamson et al. 2020). Consistent with earlier evidence for influenza and other respiratory diseases, a recent review of several studies suggests that smoking is associated with worse outcomes from COVID-19 infection (Vardavas and Nikitara 2020). Access to high-quality health care, which depends in part on health insurance coverage as well as reliable and comprehensive health information, is an additional important determinant of mortality risk in general as well as from COVID-19 (Thakur et al. 2020).

An obvious risk factor for COVID-19 is the prevalence of infection in the local community, which affects the likelihood that an individual’s day-to-day contacts with others will result in viral transmission. By modifying individual and group behaviors, local and state policies can have a substantial impact on these risks. Strong policy responses, such as lockdowns of businesses, restrictions on size of gatherings and travel, border closures, and quarantines have been associated with reduced COVID-19 transmission (Hsiang et al. 2020; Chaudhry et al. 2020).

Tribal nations appear to be especially vulnerable to COVID-19. The high rates of poverty and generally poor living conditions characterizing reservation life have been implicated in excessive rates of infectious disease and mortality among American Indians for two hundred years or more before the arrival of COVID-19 (Jones 2006).

There are currently 326 Federal Indian reservations that are sovereign nations within the U.S. (Rodriguez-Lonebear et al. 2020). Until the 1950s, most Native Americans lived on reservations, but many relocated voluntarily or were coerced to move subsequently. A large number of reservations are located in rural areas. Today, almost four-fifths of Native Americans live off the reservation, largely in metropolitan areas (U.S. Department of Health and Human Services 2018). However, this proportion varies substantially across states with, for example, over 60% of Native Americans living on reservations in South Dakota and Montana in contrast to fewer than 5% in Texas, Alaska, Florida and Colorado.(see Table 1).^2^

**Table 1:** Number of Native Americans, number of COVID-19 deaths for Native Americans, and percent of Native Americans on reservations, 16 states^a^.

| State | # of Native Americans | # of COVID-19 deaths for Native Americans | % Native Americans on reservations |
| --- | --- | --- | --- |
| Alaska | 109,751 | 70 | 1.1 |
| Arizona | 317,414 | 1,014 | 51.0 |
| California | 303,998 | 119 | 5.7 |
| Colorado | 54,847 | 51 | 4.9 |
| Florida | 59,320 | 29 | 3.1 |
| Michigan | 53,316 | 45 | 11.1 |
| Minnesota | 58,011 | 74 | 32.0 |
| Montana | 66,839 | 218 | 67.2 |
| New Mexico | 199,845 | 739 | 50.0 |
| New York | 79,512 | 38 | 9.3 |
| North Carolina | 123,952 | 43 | 5.4 |
| Oklahoma | 299,621 | 397 | NA<sup>b</sup> |
| South Dakota | 76,190 | 165 | 62.8 |
| Texas | 141,425 | 51 | 1.3 |
| Washington | 94,449 | 90 | 28.6 |
| Wisconsin | 51,392 | 74 | 32.2 |

<sup>a</sup> All numbers are based on Native Americans recorded as single-race.
<sup>b</sup> Until the Supreme Court ruling on July 9<sup>th</sup>, 2020, there were legal uncertainties as to what land in Oklahoma should be classified as reservation land.
Source: ACS annual counts of Native Americans are derived from five-year estimates for 2015-2019; Numbers of COVID-19 deaths are from CDC Weekly Updates by Select Demographic and Geographic Characteristics, updated as of January 21, 2021. The percentages of Native Americans on reservations are derived from 2015-2019 ACS estimates.

Native American reservations are affected by many of the risk factors for infectious disease described earlier. The poverty rate far exceeds that of the general population, approaching 40% for some Native groups, with low levels of education and high rates of unemployment and food insecurity on reservations compared with the general population (Sarche and Spicer 2008; Jernigan et al. 2012). Many studies have pointed to lack of indoor running water or, more generally, inadequate household plumbing (Deitz and Meehan 2019; Rodriguez-Lonebear et al. 2020), a serious concern for quality of life in general but a particularly critical risk factor for the spread of infectious diseases like COVID-19. The Navajo Nation, the largest and most populous reservation in the U.S., and one that has garnered considerable media attention due to its high rates of COVID-19 infection and mortality, has some of the highest water and plumbing insecurity in the U.S. with about 40% of families lacking running water in their homes (Deitz and Meehan 2019). In a study of reported cases of COVID-19 on reservations during the early stages of the pandemic, the proportion of homes lacking running water was more strongly associated with COVID-19 case rates than other household variables (Rodriguez-Lonebear et al. 2020).

Inadequate access to and poor quality of health services for Native Americans has been a serious problem for many years. During 2020, the problems seem to have become particularly acute because of obstacles that Native Americans faced in obtaining COVID-19 tests, and the challenges that affected treatment, hospitalization and recovery among those infected. Federally recognized tribes in the American Indian and Alaskan Native populations typically receive health care from the Indian Health Service (IHS), which has three branches: (1) provision of free health care primarily to those residing on or nearby reservations, (2) independent health services managed by the tribes but federally funded by the IHS, and (3) urban health services serving Native Americans living in urban areas (Sequist et al. 2011; Institute of Medicine 2003; Joe 2003).

The IHS and tribal health care systems are chronically underfunded. Many provider positions in the IHS remain unfilled, with vacancy rates of 25% in 2018 across the country and 30% for the Navajo Nation (U.S. Government Accountability Office 2018), and physicians report limited access to essential services as well as lack of high quality specialists, mental health services and diagnostic imaging (Sequist et al. 2011). Moreover, many Native Americans have no health insurance other than the IHS. Medicaid is underutilized in this population, and Native Americans frequently face discrimination and language barriers in non-IHS and non-tribal health facilities (Institute of Medicine 2003; Joe 2003; Cromer, Wofford, and Wyant 2019). Native Americans living on reservations (and in rural areas more generally) face especially severe challenges because of the extensive distance, travel time and cost involved in accessing health care, factors that pose a serious threat during a pandemic (Cromer, Wofford, and Wyant 2019; Groom et al. 2009; Rodriguez-Lonebear et al. 2020).

Although reservation life has encompassed many challenges during this period of high viral transmission, there are some health-related advantages, including a strong sense of community and preservation of culture that may offer strong social support along with protection from psychological distress (Huyser et al. 2018). In addition, COVID-19 policies on some reservations have been much stricter than those in the states where these reservations are located. For example, the Navajo Nation, which is located in New Mexico, Arizona, and Utah, has had some of the most aggressive lockdown and curfew policies in the U.S. since the start of the pandemic along with high testing rates for the coronavirus, and consistent, clear messaging from President Johnathan Nez about the importance of wearing masks (Navajo Nation Department of Health n.d.). Nevertheless, the analysis below suggests that these advantages may be outweighed by the many risk factors for COVID-19 infection and severity that are disproportionately present on reservations.

## 3. Data

We calculated death rates from COVID-19 from two sources. Provisional counts of deaths from COVID-19 by age group, race/ethnicity, and location (state and county) were taken from the National Center for Health Statistics (NCHS) for deaths through January 16, 2021 (we refer to these as 2020 deaths; Center for Disease Control and Prevention 2021a). Midyear population counts for 2019 for these same characteristics were obtained from the U.S. Census Bureau (2020).

We used two large, national surveys, the American Community Survey (ACS) and the Behavioral Risk Factor Surveillance System (BRFSS), to estimate potential risk factors for viral transmission and severity across states. The 2015-2019 ACS provided information on type of health insurance, the income-poverty ratio, and household information on living arrangements (whether the household is multigenerational and number of persons per room). Because the ACS asks about occupations in the past five years, we used a recent ACS (2018) to determine frontline worker status. In an effort to obtain sufficiently large sample sizes of Native Americans, particularly at the state-level, while also ensuring consistency of question wording, we pooled data from nine years of the BRFSS (2011-2019) for estimates of smoking status and the prevalence of seven health conditions: asthma, chronic obstructive pulmonary disease (COPD), kidney disease, cancer (excluding skin), heart disease, diabetes, and obesity. Sample sizes for each of the health conditions vary slightly due to different degrees of non-response.

Information on the extent to which each of the 16 states in our analysis remained open for business during the pandemic period was obtained from MultiState (Multistate 2020). In an effort to examine responses to the coronavirus, MultiState has measured the extent of lockdown of businesses and subsequent reopenings of the economy for state and local areas during much of the pandemic period.

## 4. Variables

### Racial classification and reservation status

We consider four mutually exclusive racial/ethnic groups: non-Latino Whites, non-Latino Blacks, Latinos, and non-Latino Native Americans (American Indians and Alaskan Natives). For convenience, we drop the term “non-Latino” from subsequent references to these groups. Information on race and ethnicity for COVID-19 fatalities are drawn from the death certificates, whereas data in the surveys derive from self-reports. In the ACS, where finer racial distinctions are available, we consider both Native Americans with single-race identification and, separately, those with multiple-race identification (excluding those reporting Latino ethnicity). Estimates from the BRFSS are based only on self-reported single-race Native Americans.

We used the My Tribal Area tool, which compiles 2015-2019 ACS estimates of reservation population by race, to calculate the number of single-race Native Americans residing on reservations or off-reservation trust land within the states in our analysis (U.S. Census Bureau n.d.); for simplicity, we refer to these estimates as living on a reservation.^3^ Because reservation status is not available on the microdata files from the 2015-2019 ACS obtained from IPUMS, we used homeland status as a proxy for reservation status in the state-specific estimates of risk factors. Homelands are similar to reservations,^4^ but include statistical lands in addition to legal ones; these statistical lands are intended to be “substantially meaningful” to local Native Americans (IPUMS USA n.d.). The variable in the ACS indicates whether the household is in a public use microdata area that includes any census block designated as a homeland area for American Indians, Alaska Natives, or Native Hawaiians. In July, 2020 the U.S. Supreme Court reclassified most tribal statistical land in Oklahoma as reservations. Because the My Tribal Area tool has not been updated since that time and current status estimates would not in any case be an adequate measure of exposure in the recent past, Oklahoma is excluded from estimates that require reservation status but is included in estimates based on homeland status (McGirt vs. Oklahoma 2020).

### Socioeconomic factors related to vulnerability

The measure of household crowding is determined from the number of persons and number of rooms in the household in the ACS. We use the proportion of households with more than one person per room, a standard assessment of overcrowding that has been applied in other studies (Blake, Kellerson, and Simic 2007).

Estimates of the frequency of frontline workers, which are based on data in the 2018 ACS, include only those individuals who held any job in the five-year period prior to the survey, regardless of age, and who reported their occupation. About 88% of these individuals reported a job held during the single-year prior to the ACS. Our definition of frontline workers is based on the definition offered by Dingel and Neiman (2020) and used by Blau, Koebe, and Meyerhofer (2020): occupations in which one-third or fewer workers can feasibly work from home, ascertained from responses to 15 questions in the Occupational Information Network (O*NET, n.d.).^5^

### Health-related variables

We estimated risk factors for the working age population. Because the prevalence of these factors may vary substantially by age, particularly for the health-related variables, and because the racial/ethnic groups differ substantially in the size of the older population, we restricted the age range to 18-60 for the ACS and 18-59 for the BRFSS. To further reduce potential bias, we age-standardized all health-related variables derived from the BRFSS.^6^

Each of the six illness variables is based on a question in the BRFSS asking respondents whether a medical professional ever told them that they had the particular condition. The measure of obesity is based on a calculation of BMI (BMI≥30), obtained from self-reported height and weight. A score was calculated as a count of the number of health conditions reported by the individual among these seven variables.

All estimates from the ACS and BRFSS are based on individual-level data, with the exception of the frequency of households comprising at least three generations and the measure of crowding, both of which are derived from household-level data.

### State openness

To determine the degree of state openness during the pandemic, we used scores developed by MultiState for the period between May 4, 2020 and December 30, 2020. The scores were based on assessments five days per week, ranged between 0 (full lockdown) to 100 (fully open), and were derived from 11 variables pertaining to stay-at-home orders: the definition of “essential business;” openings or operation of non-essential offices, non-essential retail, construction sites, personal care services, physical fitness businesses, bars, and venues servicing large groups; and approaches to reopening within the state, region and county (Multistate 2020).

## 5. Analytic Strategy

The analysis is primarily descriptive. We begin by comparing standardized 2020 COVID-19 mortality measures for the Native American, Black and Latino populations relative to Whites for the U.S. and for the 16 states. Next, to understand the variability across states, we compare potential risk factors across racial/ethnic groups, nationally and for the selected states. Because Native Americans who classifiy themselves with a single racial category are more likely to live on reservations than those who use multiple categories (Norris, Vines, and Hoeffel 2012), and because we anticipate that reservation status in itself may reflect higher levels of risk factors, we examine differences in potential vulnerability to the virus between single-race and multiracial Native American groups at the national level. At the state level, we directly compare risk factors between Native Americans residing on homelands and those living in other areas, as well as between Native Americans and other racial/ethnic groups. In the final stage of the analysis, we explore the relationship between potential risk factors and COVID-19 mortality across states for Native Americans.

The racial and ethnic groups in this analysis differ dramatically in their underlying age distributions. Appendix Figure 1, which presents the mean ages of the four ethnic groups in each of the 16 states, reveals that Latinos have the youngest distribution in all states, with the White population generally more than a decade older on average than the Latino population and at least several years older than the Native American population. Given that SARS-CoV-2 death rates increase steeply with age, this variability must be taken into account. Although these racial/ethnic groups also differ in their patterns of geographic residence across the U.S. (e.g., rural vs. urban and region), it is unclear whether our estimates should be standardized by place (i.e., county): an individual would be more likely to become infected (and die) if they lived in a county with a high infection rate, but controlling for county-level infection rates does not make sense if individuals in this person’s ethnic group are primarily responsible for the high rate (i.e., if they comprise a large proportion of infections in the county). The uncertain advantage of place standardization stems from the enormous variation in the concentration of each ethnic group across counties: e.g., whereas Native Americans comprise less than 1% of the population in most counties, they are the majority in 25 counties within the 16 states. As a result, we standardize all of our estimates by age only but we provide several comparisons with estimates standardized by both age and county to explore the robustness of our results. Below we describe the calculation for standardization by age and county; the procedure for standardization by age only is a straightforward modification.

Because age- and race-specific death rates from COVID-19 are not currently available by county, we use indirect standardization techniques to calculate the Standardized Mortality Ratio (SMR) for each racial/ethnic group. We follow the procedure used to examine racial/ethnic disparities in COVID-19 mortality early in the pandemic (Goldstein and Atherwood 2020). The SMR is a ratio of the observed number of COVID-19 deaths in a particular racial/ethnic group (during 2020) to the “expected” number. The expected number for each group reflects the number of deaths that would have occurred during 2020 had the group experienced the COVID-19 death rates by age and county of the entire US standard population^7^ but retained the observed population distribution by age and county of the particular racial/ethnic group. In addition to calculating the SMRs for the U.S., we obtain the corresponding numbers for the 16 states in the analysis based on age and county information for the state, using the same standard schedule as for the nationwide calculation.

In the first part of the analyis, we examine the SMR for each of the three disadvantaged groups (Native American, Black and Latino populations) relative to (i.e., divided by) the SMR for Whites. We present three sets of relative SMRs: controlling only for age, controlling only for place, and controlling for both age and place. To illustrate the importance of standardization, we also calculate the crude death rates from COVID-19 for each of these groups relative to the crude death rate for Whites. The crude death rate, so-called because it does not take age into account, is defined simply as the number of deaths from COVID-19 for a particular group relative to the total population for that group.

Analyses that examine potential risk factors for the virus across racial/ethnic groups are based on the ACS and BRFSS (the openness scores are an exception). The estimates of the prevalence of risk factors are weighted based on the sampling weights in the ACS and BRFSS, but numbers of survey participants and households shown in the tables are unweighted. In order to minimize our inclusion of unreliable estimates, state estimates are not shown when the number of households or individuals falls below 50. In estimated associations that examine only Native Americans (i.e., correlations of risk factors with COVID-19 mortality across the states), we measure COVID-19 mortality with the (age-standardized) SMR for Native Americans.

Because our dataset includes only 16 states, we restrict statistical measures to simple correlations between the SMR and risk factors rather than undertake multiple regression analysis. Confidence intervals (95%) for the age-standardized SMRs for each of the four racial/ethnic groups are presented in Appendix Figure 2 and are calculated according to the procedure presented in Ulm (1990).

## 6. Results

Figure 1 shows the crude death rates and the three sets of SMRs for COVID-19 in the U.S. during 2020 for each of the Native American, Black and Latino populations relative to Whites. These estimates reveal that, according to any of the three standardization methods, COVID-19 mortality among Native Americans relative to Whites is substantially higher than suggested by crude death rates. Based on either standardization by age or standardization by age and place, the SMR for Native Americans is 2.8 times as high as among Whites and is considerably higher than the corresponding values for Blacks and Latinos.

**Figure 1:**
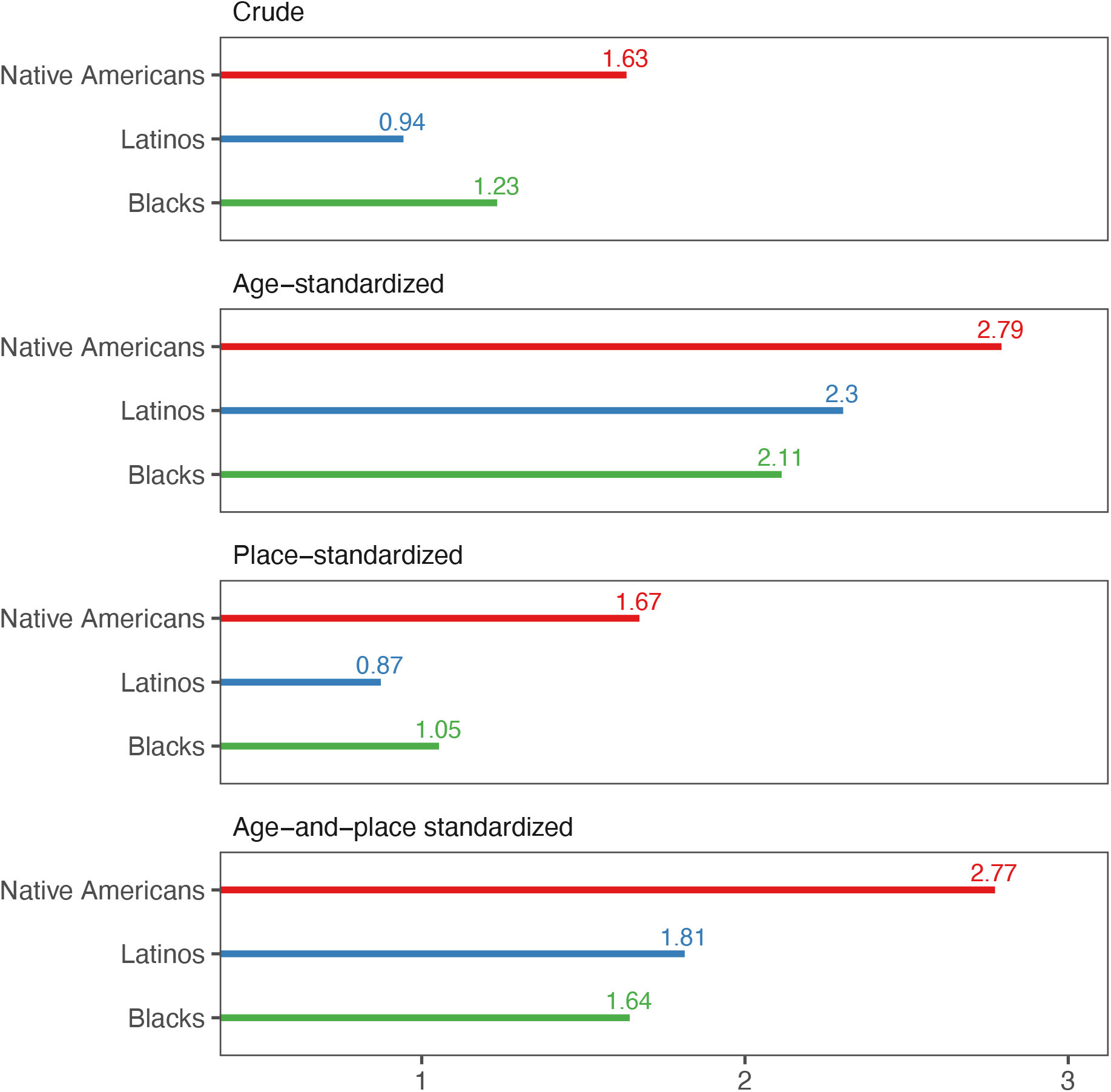
Crude Death Rates and Standardized Mortality Ratios (relative to Whites) for COVID-19 in the U.S. Source: CDC Weekly Updates by Select Demographic and Geographic Characteristics (as of Jan 21, 2021).

Figure 2 presents the age-standardized SMRs for the Black, Latino and Native American populations relative to Whites in the 16 states. The estimates underscore the huge geographic variability in COVID-19 mortality. For the Native American population, these values are about three or greater in half of the states, ranging from below one in Florida, New York and Texas to over ten in Alaska and Montana.

**Figure 2:**
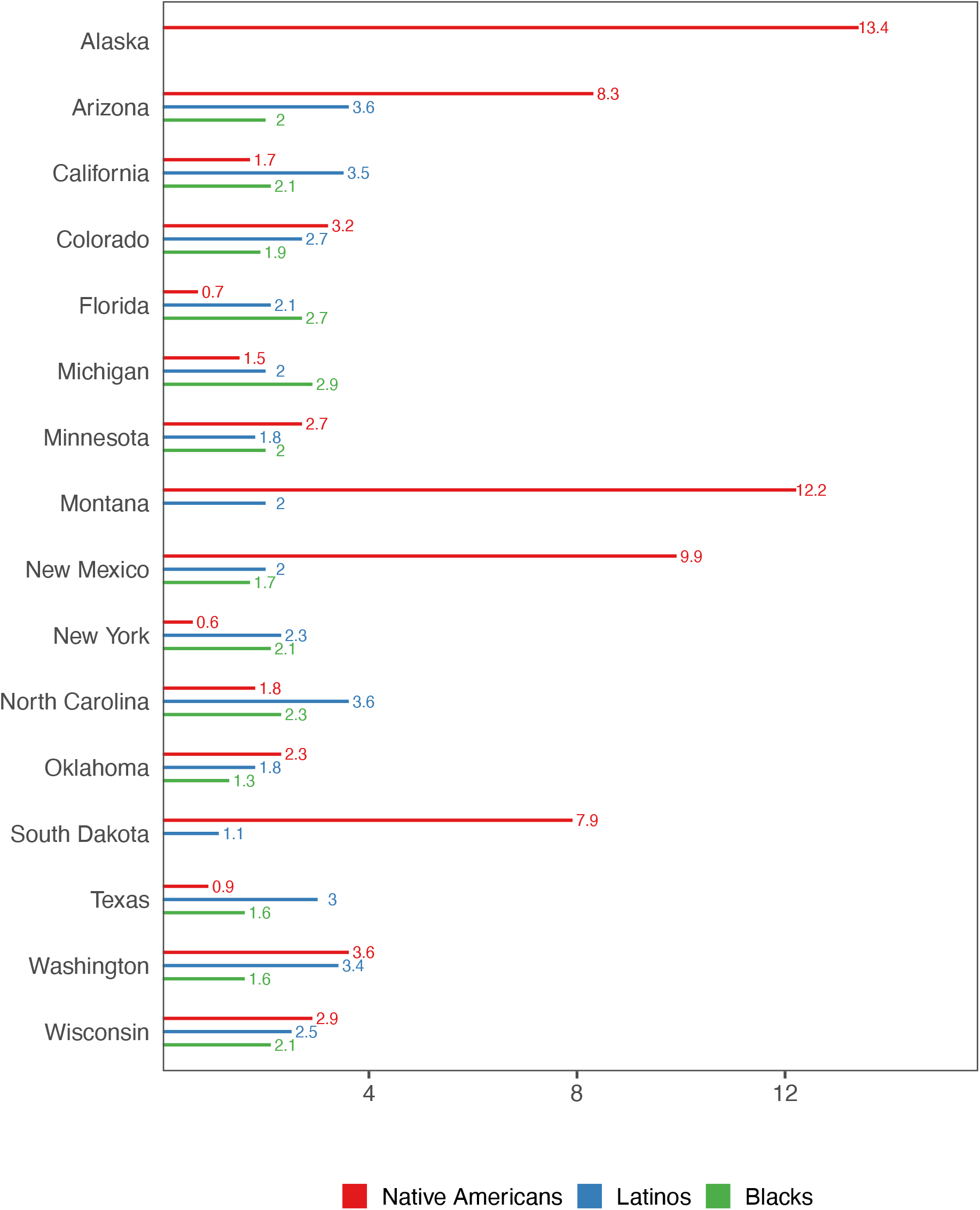
Standardized Mortality Ratios (relative to Whites) for COVID-19, 16 states. Source: CDC Weekly Updates by Select Demographic and Geographic Characteristics (as of Jan. 21, 2021). *Note: SMRs for several racial/ethnic groups are not shown because COVID-19 deaths below 10 are suppressed in the data file*.

Table 1 presents the size of the Native American population annualized from the 2015-2019 ACS, the number of COVID-19 deaths in 2020 for Native Americans, and the percentage of Native Americans living on reservations, for the 16 states. The scatterplot in Figure 3 shows the SMRs for Native Americans (no longer relative to Whites) versus the percent of Native Americans residing on reservations across the states; the size of each circle is proportional to the population of Native Americans in the state.^8^ The pattern suggests a strong correlation (r= 0.88) between living on a reservation and the SMR for COVID-19, with South Dakota and Montana having extremely large values on both of these measures in contrast, for example, to New York, Texas, Florida and California where few Native Americans reside on reservations and the SMR is relatively low. We examined the sensitivity of this correlation to the SMR controlling for both age and county, which continues to be high (r=0.77). Nevertheless, we recognize that estimates based on 16 states may not be robust and statistical testing could be misleading.

**Figure 3:**
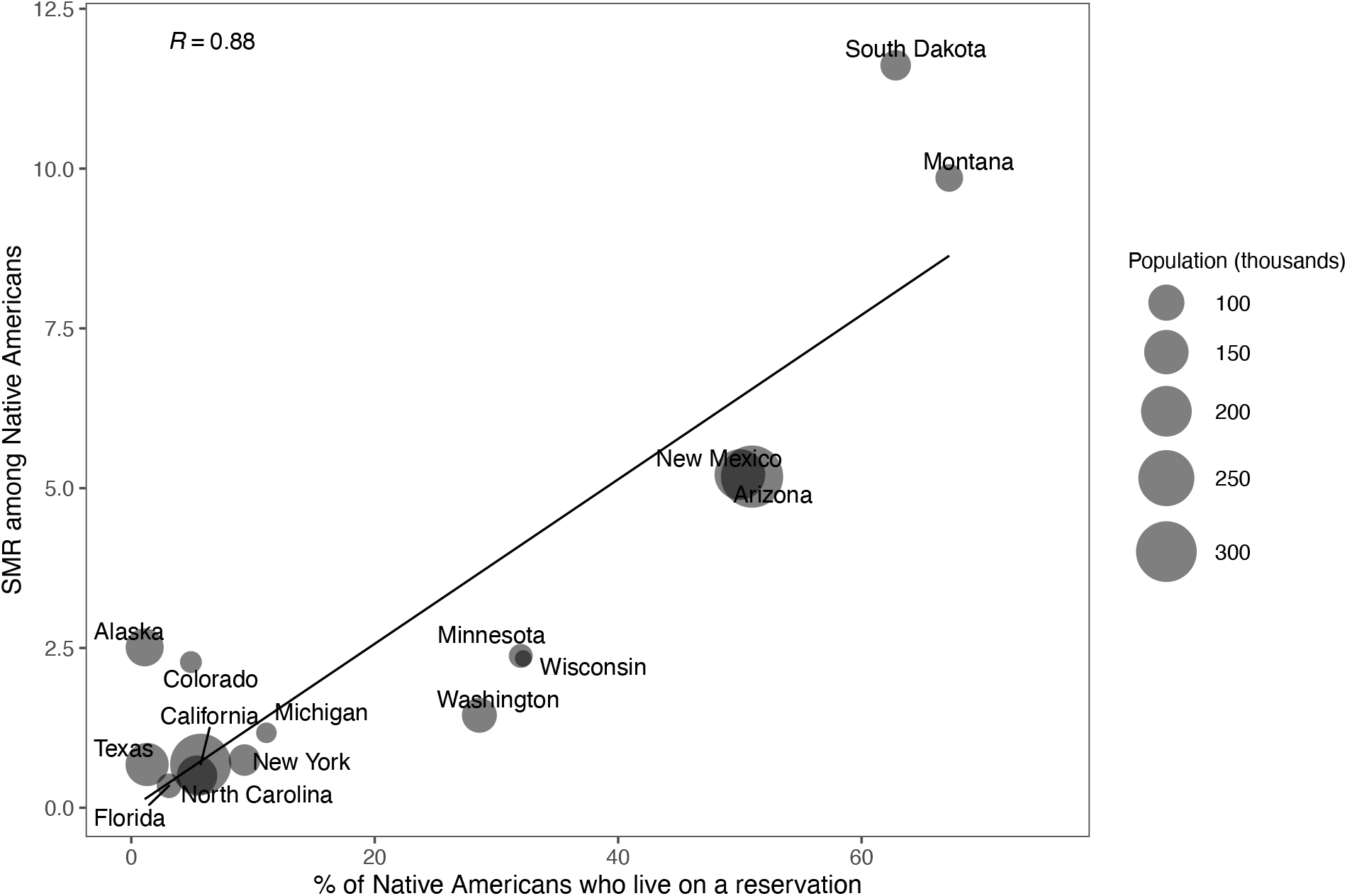
Correlation (and regression line) between the percent of Native Americans on reservations and the Standardized Mortality Ratio among Native Americans across states. Source: ACS 5**-**year estimates and CDC Weekly Updates by Select Demographic and Geographic Characteristics (as of Jan 21, 2021).

Table 2 presents the risk factors derived from the ACS and the BRFSS for the four racial/ethnic groups for the U.S. Corresponding values for the 16 states are given in Appendix Table 1. The estimates in Table 2 indicate that potential exposure to COVID-19 among Native Americans is higher than that among the other racial/ethnic groups and that single-race Native Americans appear to be at higher risk for COVID-19 exposure than their multiple-race counterparts. Single-race Native Americans have the lowest incomes, on average, and are most likely (along with Latinos) to be without health insurance (excluding the IHS). Second only to Latinos, single-race Native Americans are the most likely group to hold frontline occupations and to live in multigenerational and crowded households. The most common frontline occupations held by Native Americans for the period prior to the pandemic included cashiers, janitors and building cleaners, maids, construction and other laborers, and drivers, all relatively low paying positions that require face-to-face exposure with others (authors’ calculations). At least during most of 2020, workers in these low-income jobs were not typically provided with personal protective equipment and were likely to encounter weak safety standards in their places of employment (Carlsten et al. 2021; Gaitens et al. 2021).

**Table 2:**
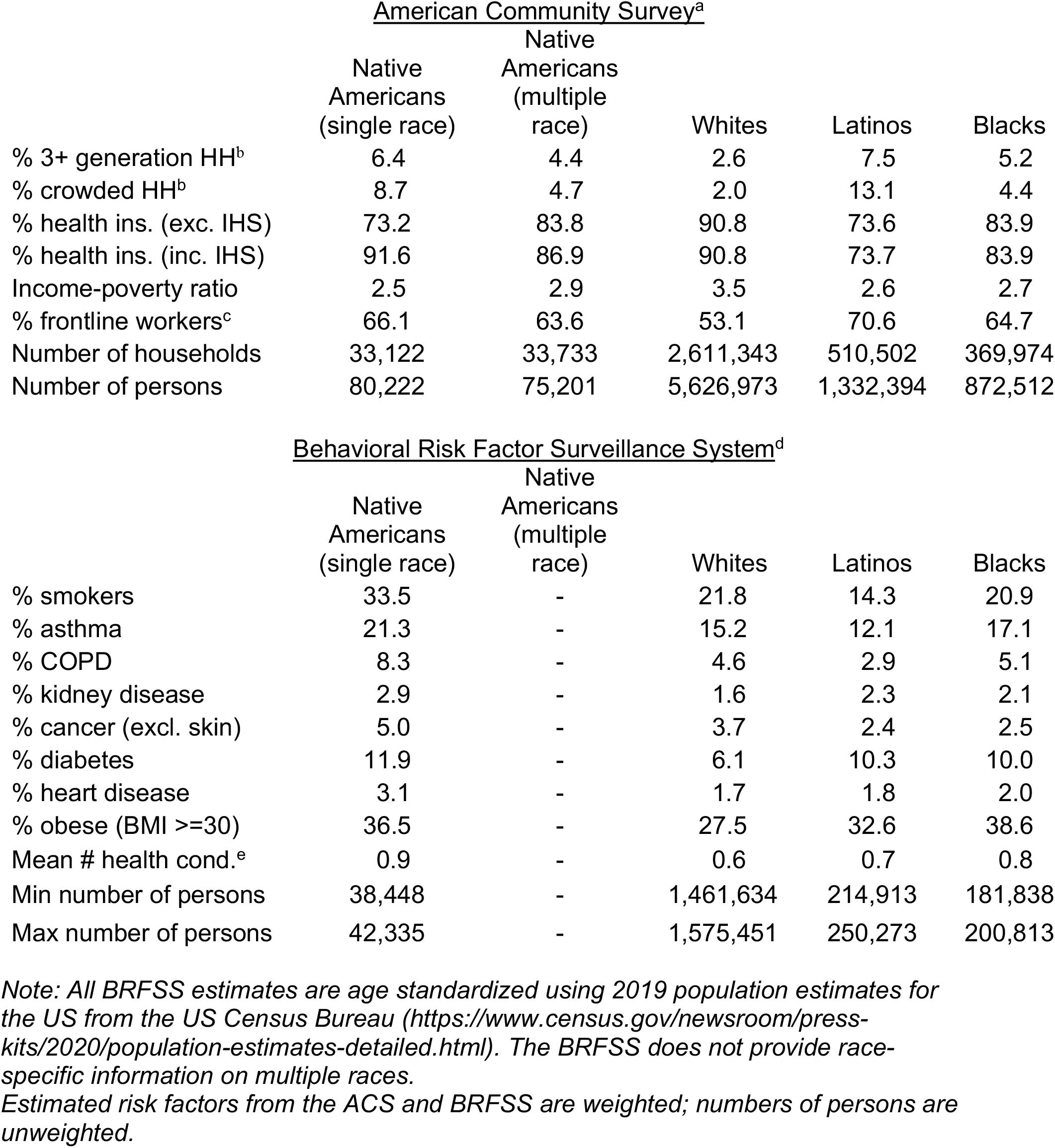
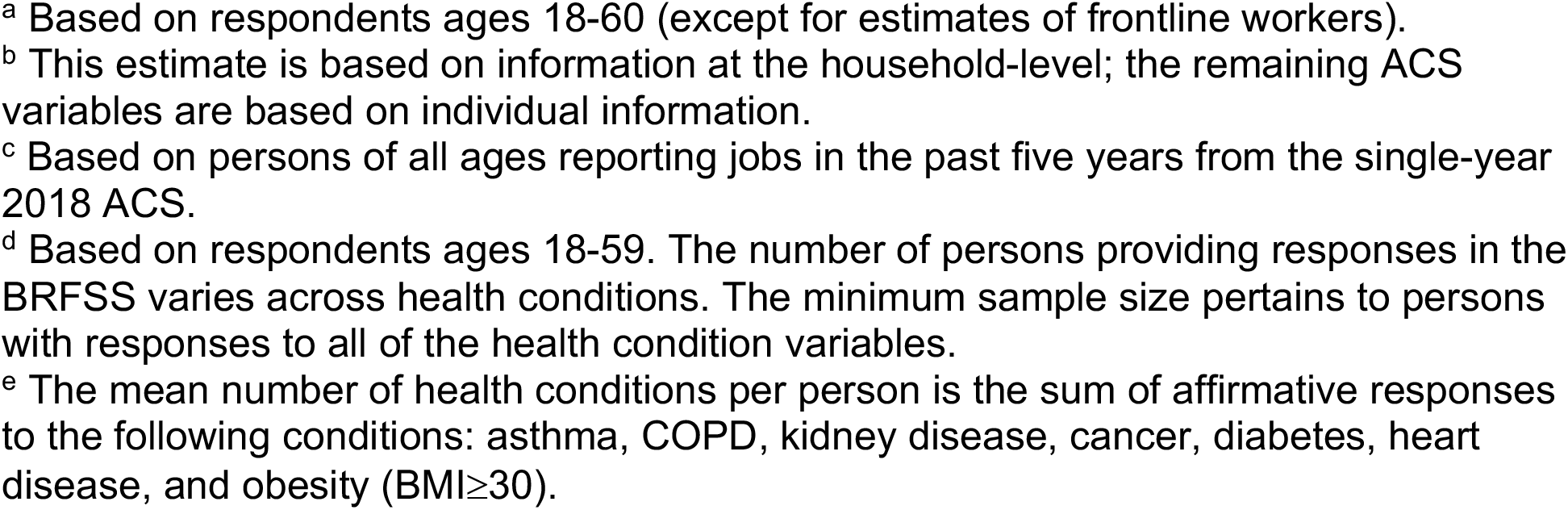
Risk factors for COVID-19 infection and severity by race and ethnicity in the U.S., American Community Survey (2015-2019) and the Behavioral Risk Factor Surveillance System (2011-2019)

| <u>American Community Survey<sup>a</sup></u> |  |  |  |  |  |
| --- | --- | --- | --- | --- | --- |
|  | Native Americans (single race) | Native Americans (multiple race) | Whites | Latinos | Blacks |
| % 3+ generation HH<sup>b</sup> | 6.4 | 4.4 | 2.6 | 7.5 | 5.2 |
| % crowded HH<sup>b</sup> | 8.7 | 4.7 | 2.0 | 13.1 | 4.4 |
| % health ins. (exc. IHS) | 73.2 | 83.8 | 90.8 | 73.6 | 83.9 |
| % health ins. (inc. IHS) | 91.6 | 86.9 | 90.8 | 73.7 | 83.9 |
| Income-poverty ratio | 2.5 | 2.9 | 3.5 | 2.6 | 2.7 |
| % frontline workers<sup>c</sup> | 66.1 | 63.6 | 53.1 | 70.6 | 64.7 |
| Number of households | 33,122 | 33,733 | 2,611,343 | 510,502 | 369,974 |
| Number of persons | 80,222 | 75,201 | 5,626,973 | 1,332,394 | 872,512 |
| <u>Behavioral Risk Factor Surveillance System<sup>d</sup></u> |  |  |  |  |  |
|  | Native Americans (single race) | Native Americans (multiple race) | Whites | Latinos | Blacks |
| % smokers | 33.5 | - | 21.8 | 14.3 | 20.9 |
| % asthma | 21.3 | - | 15.2 | 12.1 | 17.1 |
| % COPD | 8.3 | - | 4.6 | 2.9 | 5.1 |
| % kidney disease | 2.9 | - | 1.6 | 2.3 | 2.1 |
| % cancer (excl. skin) | 5.0 | - | 3.7 | 2.4 | 2.5 |
| % diabetes | 11.9 | - | 6.1 | 10.3 | 10.0 |
| % heart disease | 3.1 | - | 1.7 | 1.8 | 2.0 |
| % obese (BMI ≥30) | 36.5 | - | 27.5 | 32.6 | 38.6 |
| Mean # health cond.<sup>e</sup> | 0.9 | - | 0.6 | 0.7 | 0.8 |
| Min number of persons | 38,448 | - | 1,461,634 | 214,913 | 181,838 |
| Max number of persons | 42,335 | - | 1,575,451 | 250,273 | 200,813 |

*Note: All BRFSS estimates are age standardized using 2019 population estimates for the US from the US Census Bureau (https://www.census.gov/newsroom/press-kits/2020/population-estimates-detailed.html). The BRFSS does not provide race-specific information on multiple races.*
*Estimated risk factors from the ACS and BRFSS are weighted; numbers of persons are unweighted.*
<sup>a</sup> Based on respondents ages 18-60 (except for estimates of frontline workers).
<sup>b</sup> This estimate is based on information at the household-level; the remaining ACS variables are based on individual information.
<sup>c</sup> Based on persons of all ages reporting jobs in the past five years from the single-year 2018 ACS.
<sup>d</sup> Based on respondents ages 18-59. The number of persons providing responses in the BRFSS varies across health conditions. The minimum sample size pertains to persons with responses to all of the health condition variables.
<sup>e</sup> The mean number of health conditions per person is the sum of affirmative responses to the following conditions: asthma, COPD, kidney disease, cancer, diabetes, heart disease, and obesity (BMI ≥ 30).

Native Americans also have the poorest health profile in terms of COVID-19 susceptibility as demonstrated by the age-standardized proportions in the lower panel of Table 2. They have the highest prevalence of smoking, COPD, kidney disease, cancer, heart disease, and diabetes among the four groups and the level of obesity among Native Americans is exceeded only by the Black population. For the most part, the corresponding state-specific estimates, presented in the appendix, also show higher levels of co-morbidies among Native Americans relative to the other groups. It is quite likely that some of these estimates are too low, especially for the Native American and Latino populations, because they depend on whether the a health professional told the respondent that he/she had the condition and, thus, on access to medical care.

Figure 4 presents scatterplots between the SMRs and state-specific risk factors for Native Americans, including the average openness scores in Table 3 and eight of the risk factors in the appendix. The graphs are not presented for several health conditions with low prevalence (cancer, heart disease, COPD and kidney disease) nor for asthma, which does not have a well-established link with COVID-19 at the present time. The scatterplots reveal particularly strong correlations (≥0.77 in absolute value, Figure 5) with the SMR across states for the income-poverty ratio and the prevalence of multigenerational families and health insurance (excluding the IHS). Correlations with obesity and diabetes are also substantial (0.55 and 0.65 respectively, Figure 5). South Dakota, the state with the highest SMR, has one of the most extreme values on all of these risk factors as well as the second highest proportion of Native Americans residing on reservations.

**Figure 4:**
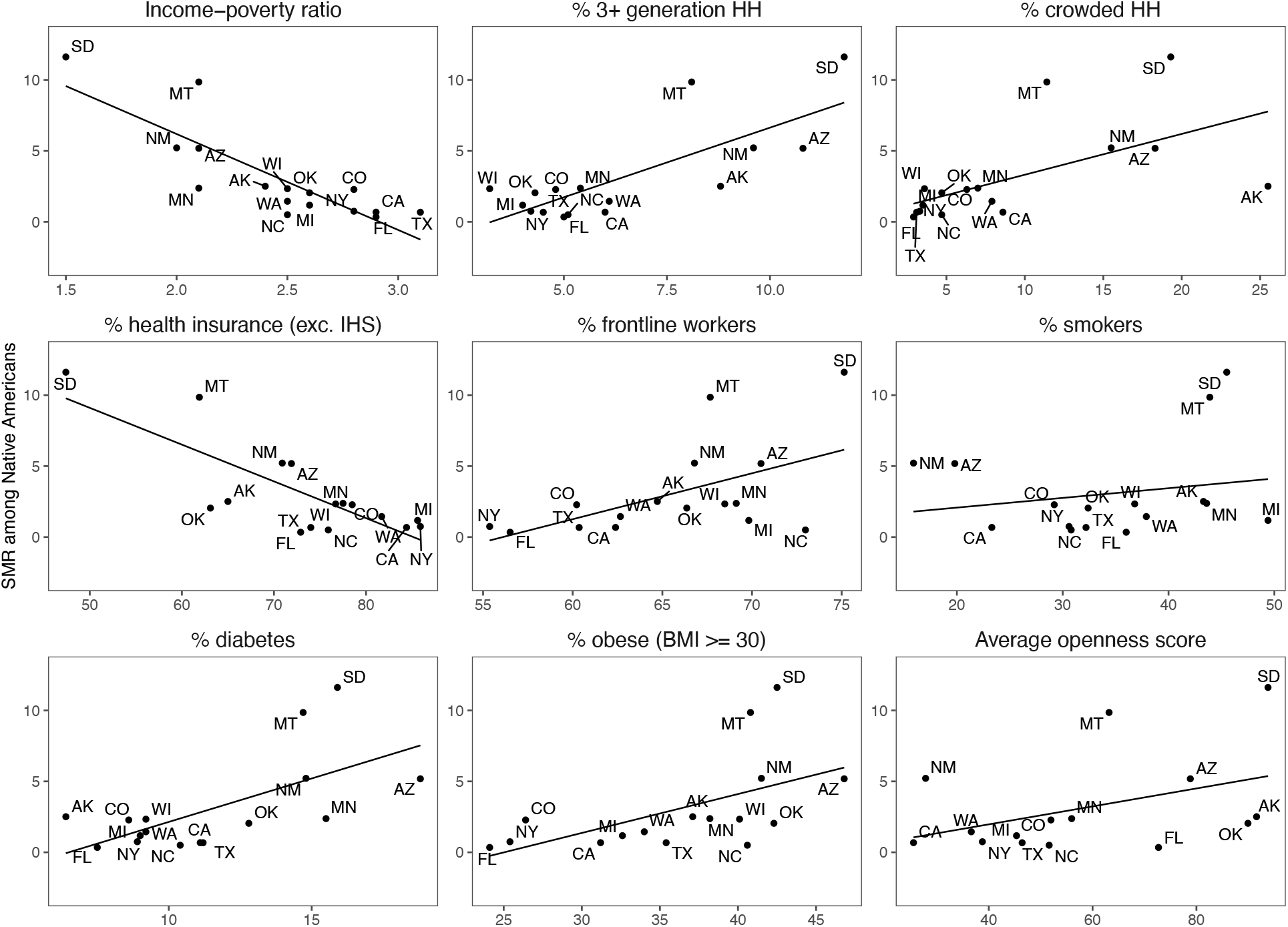
Correlations (and regression lines) between selected risk factors for COVID-19 infection and severity and the Standardized Mortality Ratio among Native Americans across states. Source: ACS 5-year estimates, ACS 1-year estimates, BRFSS, and CDC Weekly Updates by Select Demographic and Geographic Characteristics (as of Jan. 21, 2021)

**Table 3:** State openness scores, 16 states, 2020.

|  | Average openness<br>score | Minimum openness<br>score | Maximum openness<br>score |
| --- | --- | --- | --- |
| Alaska | 91.6 | 55 | 96 |
| Arizona | 78.8 | 29 | 91 |
| California | 25.5 | 10 | 33 |
| Colorado | 52.0 | 46 | 56 |
| Florida | 72.7 | 44 | 96 |
| Michigan | 45.3 | 16 | 55 |
| Minnesota | 56.0 | 25 | 67 |
| Montana | 63.2 | 55 | 72 |
| New Mexico | 27.8 | 13 | 36 |
| New York | 38.8 | 9 | 49 |
| North Carolina | 51.6 | 18 | 63 |
| Oklahoma | 89.9 | 80 | 93 |
| South Dakota | 93.8 | 92 | 96 |
| Texas | 46.4 | 29 | 52 |
| Washington | 36.6 | 13 | 46 |
| Wisconsin | 59.7 | 25 | 63 |

*Note: These scores were obtained from MultiState:*
*(https://www.multistate.us/pages/covid-19-state-reopening-guide)*
*These estimates cover the period beginning May 4<sup>th</sup>, 2020 until December 30<sup>th</sup>, 2020 and were assessed five days per week. The score evaluates the degree of state 'openness,' ranging from 0 (full lockdown) to 100 (fully open). Eleven factors are evaluated to create the score; see the text.*

**Figure 5:**
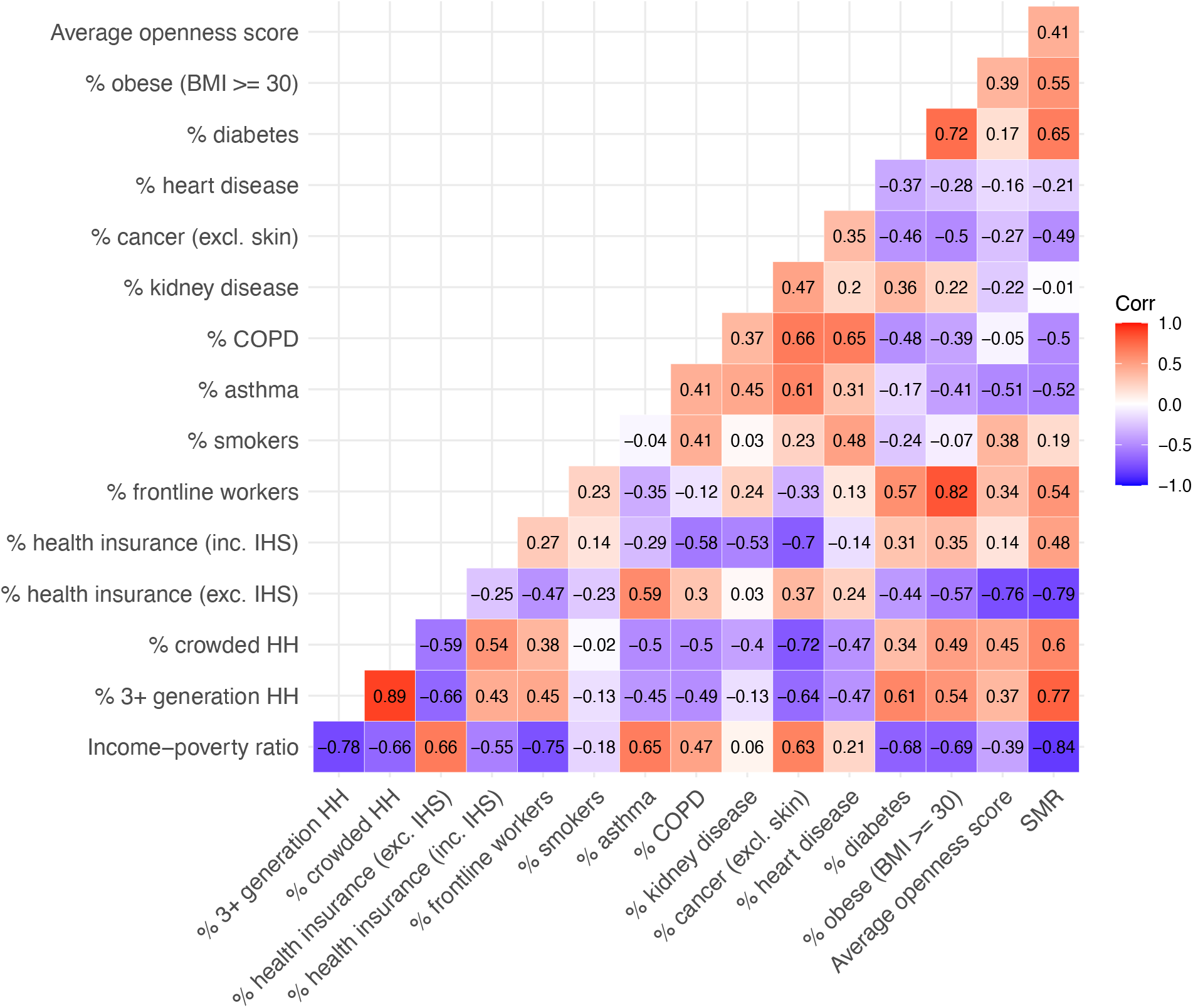
Heatmap of correlations between risk factors for COVID-19 infection and severity and the Standardized Mortality Ratio among Native Americans across states. Source: ACS 5-year estimates, ACS 1-year estimates, BRFSS, and CDC Weekly Updates by Select Demographic and Geographic Characteristics (as of Jan. 21, 2021)

As noted earlier, we do not fit multiple regresson models, both because we have only 16 observations and because many of the risk factors are highly correlated with one another, suggesting the co-occurrence of risk factors for COVID-19 and rendering it impossible to determine which are most salient. The correlations among the risk factors, as well as between the risk factors and the SMR, are shown in a heatmap in Figure 5. Not surprisingly, the income-poverty ratio is highly correlated with the presence of multigenerational households and household crowding, the proportion of Native Americans with health insurance (excluding the IHS), and the proportion with frontline occupations. These findings suggest that Native Americans are at risk of dying from COVID-19 through multiple exposures. Appendix Table 2 shows the corresponding correlations between the SMRs and risk factors for Native Americans for all-cause mortality in 2019 compared with COVID-19 mortality in 2020. The two sets of correlations are similar for many variables, emphasizing the importance of these risk factors for many causes of death in the Native American population, but the correlations for obesity, diabetes and the income-poverty ratio are larger when based on COVID-19 mortality rather than all-cause mortality.

To establish whether Native Americans living on homelands face higher risks than others, the state tables in the Appendix Table 1 stratify the ACS estimates for single-race Native Americans by homeland status. Although we cannot consider the association between risk factors and COVID-19 mortality separately by homeland status (because the COVID-19 death information does not include reservation or homeland status), these data permit us to assess the extent to which risk factors vary between those residing on vs. off homelands. For the vast majority of estimates across the states, the prevalence of COVID-19 risk factors is higher for Native Americans living on homelands than for those living off homelands. The differentials are particularly large for living conditions, i.e., multigenerational households and crowding. Not surprisingly given that IHS facilities are typically situated on or near Native lands, Native Americans on homeland areas have higher rates of health insurance when the IHS is included in the measure. However, in all states, possession of health insurance other than the IHS is more common among Native Americans residing off homelands.

## 7. Discussion

From the early days of the pandemic, the media has drawn attention to high mortality from the coronavirus among Native Americans, particularly those on select reservations. The analysis presented here measures this high fatality in a way that enhances comparability across racial and ethnic groups. Once age is taken into account, the national 2020 COVID-19 mortality level for Native Americans is almost three times that for Whites, an estimate considerably higher than the corresponding values for Blacks and Latinos. In five of the states in our analysis, Native Americans experience a mortality level from COVID-19 about eight or more times as high as that of the White population.

An examination of risk factors for COVID-19 points to several related variables that are particularly prevalent among Native Americans and that have been previously identified as risk factors for exposure to the virus or for dying among those infected. Most notable among these are high rates of poverty, which, in turn, are correlated with high density housing (high levels of crowding and multigenerational families residing in a single household), employment as frontline workers, prevalence of co-morbidites (especially obesity and diabetes) that are associated with hospitalization and mortality from the virus, and low rates of health insurance other than the IHS. It is interesting to note that the magnitude of poverty (according to the official poverty rate) among Native Americans is close to that of the Black population and only slightly higher than that of Latinos (Baker et al., 2021), yet COVID-19 mortality is considerably higher for Native American populations than for these other groups.

Health disparities between Native Americans and other groups have persisted for centuries and are reflected today in higher death rates among Native Americans for almost all major causes of death, infectious and chronic (Jones 2006). The extraordinary COVID-19 fatality rates among Native Americans are due, at least in part, to a high prevalence of illnesses and health conditions, many of which are known risk factors for COVID-19 infection and severity. In particular, extremely high rates of obesity and diabetes, which result partly from high levels of food insecurity in this population (Jernigan et al. 2013), appear to increase the risk that Native Americans acquire SARS-CoV-2 infections, develop complications and die (Zhou et al. 2020). In the current pandemic, these co-morbidites portend a poor prognosis from COVID-19.

The risk factors examined in this analysis are generally more prevalent among Native Americans who self-identify as a single-race as well as those who reside on homelands compared with their respective counterparts, underscoring potential associations between reservation life and both viral exposure and disease severity. At the same time, it is important to recognize that, although risk factors such as multigenerational households and large family compounds could exacerbate viral transmission, they also serve important social and cultural functions that could be protective of health.

Important evidence that reservation life may increase risk for COVID-19 mortality comes from the high correlation between the percent of Native Americans in a given state residing on reservations and the SMR for Native Americans in that state. This strong association suggests that large differences in the pervasiveness of reservation life may drive much of the statewide variation in COVID-19 mortality. However, we recognize that our ecological analysis cannot provide sufficient evidence of a causal connection between reservation life and the risk of COVID-19 mortality. Moreover, given that the variables in our analysis capture only some of the important components of risk, our efforts to identify the most important facets of reservation life underlying the high mortality of Native Americans remain uncertain.

A particular problem in our analysis is that possession of health insurance is a poor proxy for access to quality healthcare, a critical variable during this pandemic. Despite the establishment of treaties several generations ago, the federal government has largely neglected its obligation to provide suitable healthcare to Native Americans (Warne and Frizzel 2014). The Indian Health Service has been and continues to be woefully underfunded, a problem that is likely to be more severe among those residing on reservations who depend on the IHS for healthcare (Heisler 2016). In 2019, per capita spending for the Indian Health Service was less than half of the per capita spending on healthcare for the general U.S. population (Indian Health Service n.d.), making it extremely difficult for health facilities to acquire the resources needed to mount an effective pandemic response. For example, the IHS has few ICUs and ventilators or appropriate medical providers to treat critically ill COVID-19 patients (Schultz 2020). The historical legacies of injustice, many of which resulted from federal government actions, have created mistrust among Native Americans particularly toward the Indian Health Service and other medical institutions (Guadagnolo et al. 2009; Pacheco et al. 2013). Past experiences of discrimination, culturally and linguistically insensitive care, lack of reliable internet and phone service, and language and transportation barriers may have further reduced the likelihood that Native Americans received timely and comprehensible information about the pandemic and effective healthcare when needed (Guadagnolo et al. 2009; Joe 2003).

Another risk factor that is only partly taken into account in our analysis is inadequate housing infrastructure. Beyond the issue of crowding, Native Americans are much more likely than other groups to lack adequate plumbing in their households. About 2.5% of single-race Native American households lack hot and cold running water (in contrast to 0.2-0.4% among the other groups; authors’ calculations from the ACS). An analysis of “plumbing poverty” demonstrates that lack of complete plumbing is both spatially and racially clustered, with an especially high prevalence among Native Americans and a concentration in areas containing reservations (as high as 73 percent in one neighborhood in northern Arizona that includes five reservations; Deitz and Meehan 2019). Such water insecurity presents an obvious risk to the spread of communicable diseases and a barrier for adherence to government guidelines regarding handwashing and sanitation during the COVID-19 pandemic.

We also recognize the limitations of our measures of state openness and frontline work. The degree of openness varied over time and geographic area within the state during the pandemic and the measure does not include the extent to which residents were compliant with restrictions. Our measure of frontline work is based on pre-pandemic data and serves as a proxy for employment during the pandemic, which also varied throughout 2020 as job losses were incurred and businesses closed and reopened.

There are several additional limitations to this analysis. As mentioned previously, the lack of individual data does not permit us to assess the relative importance of the risk factors investigated here or to estimate the magnitude of risk for those residing on vs. off reservations. Furthermore, the absence of reliable data on COVID-19 cases prevents us from distinguishing the extent to which Native Americans are more susceptible than other groups to infection versus the degree to which infections in this group are more likely to result in fatalities. An additional concern relates to inaccuracies in the data on deaths from COVID-19 due to misattribution of cause of death on death certificates, exclusion of deaths indirectly attributable to COVID-19 (e.g., from delayed or foregone care from other health conditions), and errors in recording of age, race and ethnicity.

Misclassification of race and ethnicity is a particularly severe problem among Native Americans, much more so than among other groups. Previous studies have shown low levels of consistency between self-reported race on surveys and recorded race on death certificates for Native Americans and have identified substantial misclassification of Native American race on death certificates and in survey data (Espey et al. 2014; Haozous et al. 2014; Jim et al., 2014). These errors undoubtedly affect the quality of our estimates both of mortality and of risk factors obtained from the ACS and the BRFSS. Because misclassification on death certificates has generally been shown to result from underreporting of Native American race, estimates of mortality for this population are almost certainly too low (likely more so than for the other groups in this analysis), suggesting that we have underestimated the SMRs for Native Americans, as well as the SMRs relative to those for the White population. Studies have shown that the extent of underreporting varies across locations, being lower, for example, in areas with a high concentration of Native Americans than in counties and states with small Native American populations (Jim et al., 2014).^9^ Underreporting of Native American race is likely to be especially acute for those self-identifying with multiple races, who are less apt to reside in areas designated for Native American populations (Jim et al. 2014). The quality of estimates for Native Americans is further compromised by the huge variation in definitions of the Native American population across health surveys in the U.S. and the implications of such inconsistency for measurement of health disparities in the Native American population (Becker et al. 2021).

Elevated COVID-19 mortality serves as a stark reminder of the legacies of historical mistreatment of Native Americans and the continued failure of the federal, state and local governments to meet the most basic needs of this population. Our analysis shows that the death rates from COVID-19 vary substantially across states and suggests differences between those living on vs. off reservations, but there is undoubtedly further variability that we cannot assess. Previous studies have underscored heterogeneity among reservations, with substantial variability in size and, as a consequence, in the ability of tribal governments to govern effectively. There are also large differences in the prevalence of health conditions and access to healthcare (Department of the Interior n.d.; U.S. Department of Health and Human Services; Indian Health Service 2014). These disparities almost certainly have led to differential infection and fatality rates across reservations during the pandemic, above and beyond the infection rates in the broader geographic areas. Consistent with this variability, research has underscored the huge diversity among Native Americans and the unique vulnerabilities of different Native American groups (Cheek et al. 2014; Groom et al. 2009; Yellow Horse, Yang, and Huyser 2021).

Despite these critical differences in susceptibility within the Native American population, the overall set of factors associated with high risk for COVID-19 in this population are largely the same as those that have been identified as increasing the vulnerability of Native Americans to a host of diseases throughout generations, factors that reflect long-standing structural inequalities and racism. As many researchers and policy experts have noted, increased support for Native American communities is required at multiple levels, from distal to proximate variables that affect both exposure to and treatment of disease. A desperately needed first step to reduce COVID-19 fatalities among Native Americans during the ongoing pandemic, and to avert a disproportionate impact in the next one, is increased funding and coordinated policies for improvements in public health, basic infrastructure, and access to quality healthcare. At the same time, available data, although incomplete, suggest that Native Americans have had higher COVID-19 vaccination rates than the Black, Latino and White populations (Center for Disease Control and Prevention, 2020). an accomplishment that has the potential to avert disproportionately high rates of COVID-19 morbidity and mortality in the future.

## Data Availability

All data are publicly available through websites of the CDC, US Census, ACS and BRFSS.

## Acknowledgments

We would like to acknowledge financial support from the Center for Health and Wellbeing at Princeton University, assistance with the frontline calculations from Keubok Lee, helpful comments on an earlier draft from Theresa Andrasfay, Angus Deaton, Dana Glei, Gracie Himmelstein, Anne Pebley and three anonymous reviewers, and assistance with the tables from Kristen Cuzzo.

## Footnotes

1 About 95% of those reporting a single-race AI/AN for the U.S. report American Indian race rather than Alaskan Native. The only states in this analysis where the percentage of Alaskan Natives exceeds 2% are Alaska (94%), Colorado (3%), and Washington (8%).

2 This is based on Native Americans who classify themselves with a single race in the American Community Survey.

3 We provided an approximation for reservations that were in multiple states for which no reliable breakdown of state populations was available.

4 The median absolute difference in the percent of single-race Native Americans living on homeland areas vs. living on reservations across the 15 states (excluding Oklahoma) equals 0.6 percentage points.

5 O*NET is a database of work characteristics by detailed occupation collected by the U.S. Departmemt of Labor. See Goldman et al. (2021) for details of the construction of the frontline status variable.

6 The standardization is based on 2019 population estimates for the entire U.S. based on the following age groups: 18-29, 30-39, 40-49, and 50-59.

7 The standard mortality schedule is derived from COVID-19 deaths for 2020 for all races and ethnic groups combined and the entire U.S. population for 2019, with deaths and population based on 10-year age groups after the first age group of 0-4 (U.S. Census Bureau 2019).

8 While the percent of Native Americans living on reservations in Alaska is low, the majority of Native Americans in Alaska live on homelands. Most tribal lands in Alaska are considered Alaska Native Statistical Villages, which are considered homelands but not reservations. The discrepancy between living on reservations and homelands is also large for North Carolina because the majority of tribal land is considered State Designated Tribal Statistical Areas, which are homelands but not reservations. For reasons discussed earlier, Oklahoma is not included in this graph.

9 Jim et al. (2014) found lower levels of racial misclassification of Native American race in IHS Contract Health Service Delivery Areas (CHDSA), suggesting that there may be more racial awareness in these areas due to a higher proportion of Native Americans (Jim et al, 2014). All reservation lands are in CHSDA counties although not all CHSDA counties have reservations.

