## Appendix tables and figures for "COVID-19 Risk Factors and Mortality among Native Americans"

Appendix Table 1: Risk factors for COVID-19 infection and severity by race and ethnicity for 16 states: American Community Survey (2015-2019) and the Behavioral Risk Factor Surveillance System (2011-2019)

Appendix Table 2: Correlations between selected risk factors for COVID-19 infection and mortality and the Standardized Mortality Ratio among Native Americans across states

Appendix Figure 1: Average (mean) age by race and ethnicity for 16 states

Appendix Figure 2: Standardized Mortality Ratios and 95% confidence intervals for COVID-19 by race and ethnicity for 16 states

**Appendix Table 1: Risk factors for COVID-19 infection and severity by race and ethnicity for 16 states: American Community Survey (2015-2019) and the Behavioral Risk Factor Surveillance System (2011-2019)**

ALASKA

| | American Community Survey (ACS)[a] | | | | |
|---|---|---|---|---|---|
| | Native Americans (single, homeland) | Native Americans (single, not homeland) | Whites | Latinos | Blacks |
| % 3+ generation HH[b] | 8.8 | N/A | 2.3 | 4.1 | 1.7 |
| % crowded HH[b] | 25.5 | N/A | 4.3 | 5.2 | 5.5 |
| % health ins. (exc. IHS) | 65.0 | N/A | 87.2 | 81.9 | 83.3 |
| % health ins. (inc. IHS) | 97.0 | N/A | 87.6 | 83.8 | 83.9 |
| Income-poverty ratio | 2.4 | N/A | 3.7 | 3.2 | 3.1 |
| % frontline workers[c] | 64.7 | N/A | 52.7 | 60.8 | N/A |
| Number of households | 1,927 | 0[1] | 4,615 | 298 | 109 |
| Number of persons | 5,531 | 0 | 9,847 | 859 | 370 |

| | Behavioral Risk Factor Surveillance System[d] | | | | |
|---|---|---|---|---|---|
| Conditions | Native Americans (single race) | | Whites | Latinos | Blacks |
| % smokers | 43.3 | - | 18.3 | 18.6 | 22.8 |
| % asthma | 15.2 | - | 13.9 | 18.8 | 17.6 |
| % COPD | 6.2 | - | 3.2 | 3.8 | 2.9 |
| % kidney disease | 1.1 | - | 1.1 | 2.1 | 1.6 |
| % cancer (excl. skin) | 2.8 | - | 3.4 | 2.6 | 3.3 |
| % diabetes | 6.4 | - | 5.3 | 7.8 | 9.1 |
| % heart disease | 1.9 | - | 1.0 | 0.9 | 1.3 |
| % obese (BMI >=30) | 37.1 | - | 27.3 | 31.0 | 39.8 |
| Mean # health cond.[e] | 0.7 | - | 0.6 | 0.7 | 0.8 |
| Min number of persons | 3,140 | - | 12,987 | 687 | 312 |
| Max number of persons | 3,454 | - | 13,796 | 764 | 335 |

[1] Most of the tribal land in Alaska is considered Alaska Native village statistical areas or Alaska Native regional corporations, which are statistical and legal geographic entities respectively. These lands are considered homelands but not reservations. Although <1% of Alaskan Natives live on reservations, almost all Alaskan Natives live on homelands because of this distinction. This likely accounts for the zero cells for ACS estimates in Alaska.

#### ARIZONA

American Community Survey (ACS)[a]

| | Native Americans (single, homeland) | Native Americans (single, not homeland) | Whites | Latinos | Blacks |
|---|---|---|---|---|---|
| % 3+ generation HH[b] | 12.3 | 7.4 | 2.8 | 8.2 | 5.0 |
| % crowded HH[b] | 19.5 | 15.8 | 2.6 | 12.3 | 6.7 |
| % health ins. (exc. IHS) | 70.7 | 75.3 | 90.2 | 74.9 | 85.9 |
| % health ins. (inc. IHS) | 93.6 | 93.3 | 90.2 | 75.3 | 85.9 |
| Income-poverty ratio | 1.9 | 2.5 | 3.4 | 2.5 | 2.7 |
| % frontline workers[c] | 71.3 | 68.8 | 51.3 | 69.3 | 59.5 |
| Number of households | 3,858 | 641 | 47,622 | 19,051 | 2,950 |
| Number of persons | 10,184 | 1,747 | 100,185 | 49,708 | 7,120 |

Behavioral Risk Factor Surveillance System[d]

| Conditions | Native Americans (single race) | | Whites | Latinos | Blacks |
|---|---|---|---|---|---|
| % smokers | 19.8 | - | 20.0 | 14.6 | 18.7 |
| % asthma | 19.4 | - | 17.8 | 11.9 | 17.5 |
| % COPD | 3.9 | - | 4.5 | 3.6 | 5.4 |
| % kidney disease | 3.0 | - | 1.9 | 2.8 | 2.6 |
| % cancer (excl. skin) | 2.8 | - | 3.6 | 2.7 | 2.5 |
| % diabetes | 18.8 | - | 5.7 | 10.5 | 9.9 |
| % heart disease | 0.9 | - | 1.7 | 1.8 | 0.9 |
| % obese (BMI >=30) | 46.8 | - | 24.7 | 35.7 | 33.5 |
| Mean # health cond.[e] | 1.0 | - | 0.6 | 0.7 | 0.7 |
| Min number of persons | 2,318 | - | 21,865 | 7,443 | 1,129 |
| Max number of persons | 2,589 | - | 23,597 | 8,534 | 1,261 |

### CALIFORNIA

American Community Survey (ACS)[a]

| | Native Americans (single, homeland) | Native Americans (single, not homeland) | Whites | Latinos | Blacks |
|---|---|---|---|---|---|
| % 3+ generation HH[b] | 7.8 | 5.2 | 2.6 | 9.8 | 5.1 |
| % crowded HH[b] | 10.1 | 7.8 | 3.3 | 21.4 | 6.7 |
| % health ins. (exc. IHS) | 80.2 | 86.7 | 93.8 | 81.7 | 90.6 |
| % health ins. (inc. IHS) | 94.7 | 91.8 | 93.8 | 81.8 | 90.6 |
| Income-poverty ratio | 2.4 | 3.1 | 3.7 | 2.7 | 2.9 |
| % frontline workers[c] | 66.9 | 59.6 | 45.1 | 71.3 | 56.5 |
| Number of households | 1,313 | 983 | 190,629 | 134,463 | 20,295 |
| Number of persons | 2,924 | 2,510 | 416,817 | 387,302 | 52,303 |

Behavioral Risk Factor Surveillance System[d]

| Conditions | Native Americans (single race) | | Whites | Latinos | Blacks |
|---|---|---|---|---|---|
| % smokers | 23.3 | - | 16.2 | 11.7 | 18.4 |
| % asthma | 29.6 | - | 16.9 | 10.7 | 21.5 |
| % COPD | 5.5 | - | 3.5 | 2.4 | 4.4 |
| % kidney disease | 3.0 | - | 1.6 | 2.0 | 1.8 |
| % cancer (excl. skin) | 5.0 | - | 3.5 | 2.0 | 3.0 |
| % diabetes | 11.1 | - | 4.9 | 11.4 | 9.5 |
| % heart disease | 2.6 | - | 1.2 | 1.5 | 1.9 |
| % obese (BMI >=30) | 31.2 | - | 22.1 | 32.2 | 33.2 |
| Mean # health cond.[e] | 0.9 | - | 0.5 | 0.6 | 0.8 |
| Min number of persons | 502 | - | 28,513 | 21,892 | 3,016 |
| Max number of persons | 565 | - | 30,699 | 25,690 | 3,359 |

#### COLORADO

American Community Survey (ACS)[a]

| | Native Americans (single, homeland) | Native Americans (single, not homeland) | Whites | Latinos | Blacks |
|---|---|---|---|---|---|
| % 3+ generation HH[b] | 7.5 | 4.3 | 2.1 | 5.8 | 4.5 |
| % crowded HH[b] | 3.8 | 6.9 | 1.6 | 9.9 | 6.1 |
| % health ins. (exc. IHS) | 62.2 | 82.5 | 92.3 | 77.4 | 88.2 |
| % health ins. (inc. IHS) | 94.0 | 87.5 | 92.3 | 77.4 | 88.2 |
| Income-poverty ratio | 2.5 | 2.9 | 3.7 | 2.8 | 2.8 |
| % frontline workers[c] | N/A | 61.0 | 47.5 | 69.9 | 53.9 |
| Number of households | 121 | 271 | 55,763 | 10,318 | 1,857 |
| Number of persons | 270 | 676 | 115,238 | 25,523 | 4,634 |

Behavioral Risk Factor Surveillance System[d]

| Conditions | Native Americans (single race) | | Whites | Latinos | Blacks |
|---|---|---|---|---|---|
| % smokers | 29.2 | - | 17.7 | 17.8 | 24.1 |
| % asthma | 20.4 | - | 15.3 | 11.2 | 17.1 |
| % COPD | 4.9 | - | 2.8 | 2.2 | 3.4 |
| % kidney disease | 1.7 | - | 1.2 | 1.7 | 1.9 |
| % cancer (excl. skin) | 5.7 | - | 3.3 | 2.5 | 2.9 |
| % diabetes | 8.6 | - | 3.6 | 9.0 | 7.4 |
| % heart disease | 2.9 | - | 0.8 | 1.4 | 1.5 |
| % obese (BMI >=30) | 26.4 | - | 19.2 | 28.6 | 29.5 |
| Mean # health cond.[e] | 0.7 | - | 0.5 | 0.6 | 0.6 |
| Min number of persons | 498 | - | 41,037 | 9,159 | 1,515 |
| Max number of persons | 553 | - | 43,786 | 10,563 | 1,692 |

FLORIDA

| | American Community Survey (ACS)[a] | | | | |
|---|---|---|---|---|---|
| | Native Americans (single, homeland) | Native Americans (single, not homeland) | Whites | Latinos | Blacks |
| % 3+ generation HH[b] | 5.5 | 4.9 | 2.9 | 7.2 | 6.0 |
| % crowded HH[b] | 3.7 | 2.8 | 2.0 | 8.0 | 6.4 |
| % health ins. (exc. IHS) | 50.9 | 75.6 | 84.1 | 72.6 | 76.9 |
| % health ins. (inc. IHS) | 88.3 | 80.2 | 84.1 | 72.7 | 76.9 |
| Income-poverty ratio | 3.4 | 2.9 | 3.3 | 2.7 | 2.5 |
| % frontline workers[c] | N/A | 59.1 | 53.9 | 65.3 | 65.9 |
| Number of households | 143 | 417 | 138,306 | 51,857 | 28,550 |
| Number of persons | 247 | 953 | 293,980 | 124,578 | 70,063 |

| | Behavioral Risk Factor Surveillance System[d] | | | | |
|---|---|---|---|---|---|
| Conditions | Native Americans (single race) | | Whites | Latinos | Blacks |
| % smokers | 36.0 | - | 23.5 | 14.5 | 15.0 |
| % asthma | 20.1 | - | 13.6 | 13.8 | 13.2 |
| % COPD | 9.6 | - | 5.6 | 3.5 | 4.3 |
| % kidney disease | 2.8 | - | 1.9 | 2.5 | 1.9 |
| % cancer (excl. skin) | 7.1 | - | 3.7 | 2.9 | 2.2 |
| % diabetes | 7.5 | - | 6.2 | 7.9 | 9.1 |
| % heart disease | 1.8 | - | 2.0 | 1.9 | 2.2 |
| % obese (BMI >=30) | 24.1 | - | 24.8 | 28.1 | 35.7 |
| Mean # health cond.[e] | 0.7 | - | 0.6 | 0.6 | 0.7 |
| Min number of persons | 838 | - | 46,127 | 10,589 | 8,479 |
| Max number of persons | 926 | - | 50,233 | 13,096 | 9,515 |

<u>MICHIGAN</u>

American Community Survey (ACS)[a]

| | Native Americans (single, homeland) | Native Americans (single, not homeland) | Whites | Latinos | Blacks |
|---|---|---|---|---|---|
| % 3+ generation HH[b] | 4.6 | 3.5 | 2.4 | 4.6 | 4.8 |
| % crowded HH[b] | 4.5 | 2.6 | 1.9 | 6.8 | 3.1 |
| % health ins. (exc. IHS) | 84.3 | 86.8 | 92.7 | 81.1 | 88.8 |
| % health ins. (inc. IHS) | 95.7 | 92.5 | 92.7 | 81.2 | 88.8 |
| Income-poverty ratio | 2.5 | 2.7 | 3.3 | 2.6 | 2.3 |
| % frontline workers[c] | 68.2 | 72.0 | 58.6 | 69.9 | 68.1 |
| Number of households | 487 | 289 | 98,626 | 4,073 | 10,794 |
| Number of persons | 1,048 | 661 | 213,309 | 10,409 | 24,813 |

Behavioral Risk Factor Surveillance System[d]

| Conditions | Native Americans (single race) | | Whites | Latinos | Blacks |
|---|---|---|---|---|---|
| % smokers | 49.4 | - | 24.6 | 23.8 | 26.5 |
| % asthma | 25.4 | - | 16.8 | 16.3 | 20.7 |
| % COPD | 11.0 | - | 5.3 | 5.6 | 8.2 |
| % kidney disease | 3.0 | - | 2.0 | 1.6 | 3.1 |
| % cancer (excl. skin) | 5.7 | - | 3.9 | 3.3 | 2.5 |
| % diabetes | 9.0 | - | 6.3 | 11.3 | 9.3 |
| % heart disease | 5.0 | - | 1.8 | 3.0 | 2.1 |
| % obese (BMI >=30) | 32.6 | - | 30.0 | 39.9 | 40.7 |
| Mean # health cond.[e] | 0.9 | - | 0.7 | 0.8 | 0.9 |
| Min number of persons | 539 | - | 36,978 | 1,656 | 5,605 |
| Max number of persons | 589 | - | 39,665 | 1.825 | 6,139 |

#### MINNESOTA

American Community Survey (ACS)[a]

| | Native Americans (single, homeland) | Native Americans (single, not homeland) | Whites | Latinos | Blacks |
|---|---|---|---|---|---|
| % 3+ generation HH[b] | 6.2 | 4.0 | 1.4 | 5.2 | 3.4 |
| % crowded HH[b] | 6.8 | 7.2 | 1.5 | 11.9 | 11.5 |
| % health ins. (exc. IHS) | 74.4 | 82.9 | 95.5 | 74.4 | 89.1 |
| % health ins. (inc. IHS) | 94.7 | 90.8 | 95.5 | 74.5 | 89.1 |
| Income-poverty ratio | 2.0 | 2.3 | 3.7 | 2.5 | 2.4 |
| % frontline workers[c] | 76.1 | 50.3 | 53.4 | 71.8 | 70.1 |
| Number of households | 660 | 132 | 60,135 | 1,788 | 1,804 |
| Number of persons | 1,471 | 392 | 126,311 | 4,738 | 4,636 |

Behavioral Risk Factor Surveillance System[d]

| Conditions | Native Americans (single race) | | Whites | Latinos | Blacks |
|---|---|---|---|---|---|
| % smokers | 43.6 | - | 18.9 | 15.8 | 22.8 |
| % asthma | 20.9 | - | 12.5 | 10.3 | 15.0 |
| % COPD | 5.8 | - | 2.6 | 1.9 | 3.4 |
| % kidney disease | 3.5 | - | 1.1 | 2.2 | 2.5 |
| % cancer (excl. skin) | 4.6 | - | 3.1 | 2.4 | 1.7 |
| % diabetes | 15.5 | - | 5.0 | 11.6 | 9.6 |
| % heart disease | 2.2 | - | 1.1 | 1.7 | 1.1 |
| % obese (BMI >=30) | 38.2 | - | 26.1 | 33.7 | 31.6 |
| Mean # health cond.[e] | 0.9 | - | 0.5 | 0.6 | 0.7 |
| Min number of persons | 1,033 | - | 65,642 | 2,946 | 3,045 |
| Max number of persons | 1,144 | - | 70,625 | 3,902 | 3,464 |

### MONTANA

American Community Survey (ACS)[a]

| | Native Americans (single, homeland) | Native Americans (single, not homeland) | Whites | Latinos | Blacks |
|---|---|---|---|---|---|
| % 3+ generation HH[b] | 8.8 | N/A | 1.5 | 2.0 | N/A |
| % crowded HH[b] | 12.3 | N/A | 2.4 | 7.9 | N/A |
| % health ins. (exc. IHS) | 59.3 | 78.9 | 88.7 | 81.5 | 93.5 |
| % health ins. (inc. IHS) | 96.5 | 90.8 | 88.7 | 82.8 | 93.8 |
| Income-poverty ratio | 2.0 | 2.6 | 3.1 | 2.7 | 2.3 |
| % frontline workers[c] | 67.3 | N/A | 59.0 | 56.2 | N/A |
| Number of households | 698 | 48 | 10,683 | 278 | 35 |
| Number of persons | 1,740 | 174 | 22,035 | 716 | 110 |

Behavioral Risk Factor Surveillance System[d]

| Conditions | Native Americans (single race) | | Whites | Latinos | Blacks |
|---|---|---|---|---|---|
| % smokers | 43.9 | - | 20.4 | 30.0 | 31.6 |
| % asthma | 18.0 | - | 14.1 | 17.7 | 15.7 |
| % COPD | 5.8 | - | 3.7 | 5.0 | 5.1 |
| % kidney disease | 3.2 | - | 1.4 | 1.6 | 4.4 |
| % cancer (excl. skin) | 4.7 | - | 3.9 | 2.4 | 4.9 |
| % diabetes | 14.7 | - | 4.6 | 5.3 | 10.3 |
| % heart disease | 2.2 | - | 1.2 | 1.6 | 0.0 |
| % obese (BMI >=30) | 40.8 | - | 23.9 | 30.0 | 29.0 |
| Mean # health cond.[e] | 0.9 | - | 0.5 | 0.6 | 0.6 |
| Min number of persons | 3,224 | - | 26,213 | 810 | 100 |
| Max number of persons | 3,511 | - | 28,060 | 894 | 109 |

#### NEW MEXICO

American Community Survey (ACS)[a]

| | Native Americans (single, homeland) | Native Americans (single, not homeland) | Whites | Latinos | Blacks |
|---|---|---|---|---|---|
| % 3+ generation HH[b] | 10.8 | 4.5 | 2.3 | 5.2 | 2.0 |
| % crowded HH[b] | 17.3 | 8.1 | 1.8 | 5.9 | 2.8 |
| % health ins. (exc. IHS) | 70.1 | 75.4 | 91.3 | 82.6 | 88.0 |
| % health ins. (inc. IHS) | 97.8 | 91.2 | 91.4 | 82.9 | 88.0 |
| Income-poverty ratio | 2.0 | 2.3 | 3.2 | 2.5 | 2.6 |
| % frontline workers[c] | 70.2 | 49.6 | 49.7 | 64.4 | 67.9 |
| Number of households | 3,122 | 191 | 9,082 | 9,083 | 318 |
| Number of persons | 8,256 | 453 | 18,006 | 20,637 | 837 |

Behavioral Risk Factor Surveillance System[d]

| Conditions | Native Americans (single race) | | Whites | Latinos | Blacks |
|---|---|---|---|---|---|
| % smokers | 15.9 | - | 22.6 | 20.2 | 29.1 |
| % asthma | 14.4 | - | 17.7 | 14.0 | 16.1 |
| % COPD | 2.0 | - | 4.4 | 3.5 | 4.8 |
| % kidney disease | 2.4 | - | 1.7 | 2.3 | 3.9 |
| % cancer (excl. skin) | 2.5 | - | 4.1 | 3.3 | 2.9 |
| % diabetes | 14.8 | - | 4.8 | 9.9 | 9.5 |
| % heart disease | 1.3 | - | 1.4 | 1.4 | 3.4 |
| % obese (BMI >=30) | 41.5 | - | 24.8 | 33.1 | 35.0 |
| Mean # health cond.[e] | 0.8 | - | 0.6 | 0.7 | 0.8 |
| Min number of persons | 3,764 | - | 14,532 | 13,733 | 449 |
| Max number of persons | 4,115 | - | 15,402 | 15,122 | 479 |

NEW YORK

American Community Survey (ACS)[a]

| | Native Americans (single, homeland) | Native Americans (single, not homeland) | Whites | Latinos | Blacks |
|---|---|---|---|---|---|
| % 3+ generation HH[b] | 2.7 | 4.7 | 2.4 | 7.9 | 6.8 |
| % crowded HH[b] | 0.6 | 4.1 | 3.0 | 14.8 | 7.7 |
| % health ins. (exc. IHS) | 81.9 | 87.2 | 94.8 | 82.9 | 90.2 |
| % health ins. (inc. IHS) | 94.8 | 91.5 | 94.8 | 82.9 | 90.2 |
| Income-poverty ratio | 2.4 | 2.9 | 3.7 | 2.7 | 2.9 |
| % frontline workers[c] | 61.8 | 53.6 | 48.3 | 68.7 | 63.1 |
| Number of households | 219 | 343 | 150,438 | 30,289 | 24,829 |
| Number of persons | 496 | 846 | 335,743 | 81,031 | 66,386 |

Behavioral Risk Factor Surveillance System[d]

| Conditions | Native Americans (single race) | | Whites | Latinos | Blacks |
|---|---|---|---|---|---|
| % smokers | 30.6 | - | 19.4 | 14.7 | 17.5 |
| % asthma | 19.5 | - | 15.4 | 16.5 | 17.7 |
| % COPD | 7.2 | - | 3.8 | 3.2 | 4.4 |
| % kidney disease | 2.4 | - | 1.2 | 2.3 | 1.6 |
| % cancer (excl. skin) | 3.4 | - | 3.6 | 2.3 | 2.0 |
| % diabetes | 8.9 | - | 4.9 | 9.5 | 9.2 |
| % heart disease | 3.4 | - | 1.4 | 1.9 | 1.7 |
| % obese (BMI >=30) | 25.4 | - | 23.9 | 29.0 | 32.3 |
| Mean # health cond.[e] | 0.7 | - | 0.5 | 0.7 | 0.7 |
| Min number of persons | 579 | - | 45,905 | 9,408 | 6,743 |
| Max number of persons | 667 | - | 50,400 | 11,046 | 7,595 |

NORTH CAROLINA

American Community Survey (ACS)[a]

| | Native Americans (single, homeland) | Native Americans (single, not homeland) | Whites | Latinos | Blacks |
|---|---|---|---|---|---|
| % 3+ generation HH[b] | 5.5 | 3.9 | 2.5 | 4.6 | 5.1 |
| % crowded HH[b] | 4.6 | 5.2 | 1.7 | 11.7 | 3.4 |
| % health ins. (exc. IHS) | 74.2 | 81.1 | 87.8 | 55.4 | 82.9 |
| % health ins. (inc. IHS) | 78.3 | 82.3 | 87.8 | 55.5 | 82.9 |
| Income-poverty ratio | 2.4 | 2.7 | 3.3 | 2.2 | 2.5 |
| % frontline workers[c] | 74.5 | 67.8 | 54.6 | 77.0 | 66.7 |
| Number of households | 1,431 | 272 | 86,529 | 8,469 | 22,471 |
| Number of persons | 3,064 | 712 | 183,716 | 20,621 | 50,218 |

Behavioral Risk Factor Surveillance System[d]

| Conditions | Native Americans (single race) | | Whites | Latinos | Blacks |
|---|---|---|---|---|---|
| % smokers | 30.8 | - | 23.6 | 11.9 | 22.9 |
| % asthma | 18.5 | - | 13.9 | 7.6 | 15.0 |
| % COPD | 11.2 | - | 5.4 | 2.1 | 4.7 |
| % kidney disease | 4.1 | - | 1.8 | 2.9 | 2.4 |
| % cancer (excl. skin) | 5.9 | - | 3.6 | 1.8 | 2.6 |
| % diabetes | 10.4 | - | 6.2 | 9.9 | 10.8 |
| % heart disease | 3.8 | - | 1.9 | 1.6 | 1.9 |
| % obese (BMI >=30) | 40.6 | - | 28.0 | 30.3 | 41.9 |
| Mean # health cond.[e] | 0.9 | - | 0.6 | 0.6 | 0.8 |
| Min number of persons | 962 | - | 21,910 | 2,939 | 7,037 |
| Max number of persons | 1,050 | - | 23,591 | 3,852 | 7,795 |

OKLAHOMA

American Community Survey (ACS)[a]

| | Native Americans (single, homeland) | Native Americans (single, not homeland) | Whites | Latinos | Blacks |
|---|---|---|---|---|---|
| % 3+ generation HH[b] | 4.5 | 0.3 | 2.9 | 4.8 | 3.6 |
| % crowded HH[b] | 4.7 | 3.1 | 2.7 | 9.5 | 4.2 |
| % health ins. (exc. IHS) | 62.9 | 68.7 | 83.8 | 59.9 | 74.2 |
| % health ins. (inc. IHS) | 96.0 | 94.7 | 84.3 | 61.2 | 74.6 |
| Income-poverty ratio | 2.6 | 2.7 | 3.1 | 2.3 | 2.4 |
| % frontline workers[c] | 66.1 | N/A | 56.5 | 77.7 | 66.2 |
| Number of households | 4,119 | 87 | 31,462 | 3,449 | 2,583 |
| Number of persons | 9,315 | 194 | 66,601 | 8,615 | 5,961 |

Behavioral Risk Factor Surveillance System[d]

| Conditions | Native Americans (single race) | | Whites | Latinos | Blacks |
|---|---|---|---|---|---|
| % smokers | 32.4 | - | 25.2 | 17.9 | 24.0 |
| % asthma | 19.2 | - | 15.7 | 9.8 | 18.8 |
| % COPD | 6.7 | - | 5.8 | 3.0 | 6.2 |
| % kidney disease | 2.5 | - | 1.7 | 2.7 | 2.5 |
| % cancer (excl. skin) | 4.3 | - | 3.6 | 2.7 | 3.6 |
| % diabetes | 12.8 | - | 7.2 | 11.5 | 10.2 |
| % heart disease | 3.0 | - | 2.2 | 2.0 | 2.1 |
| % obese (BMI >=30) | 42.3 | - | 33.3 | 35.9 | 39.9 |
| Mean # health cond.[e] | 0.9 | - | 0.7 | 0.7 | 0.8 |
| Min number of persons | 2,460 | - | 21,293 | 2,301 | 2,041 |
| Max number of persons | 2,664 | - | 22,857 | 2,813 | 2,229 |

SOUTH DAKOTA

American Community Survey (ACS)[a]

| | Native Americans (single, homeland) | Native Americans (single, not homeland) | Whites | Latinos | Blacks |
|---|---|---|---|---|---|
| % 3+ generation HH[b] | 13.1 | N//A | 1.1 | 2.7 | 5.0 |
| % crowded HH[b] | 19.3 | NA | 1.2 | 7.0 | 6.5 |
| % health ins. (exc. IHS) | 46.9 | 53.4 | 90.5 | 72.0 | 77.2 |
| % health ins. (inc. IHS) | 97.0 | 83.2 | 90.6 | 73.7 | 77.2 |
| Income-poverty ratio | 1.5 | 1.6 | 3.4 | 2.4 | 2.3 |
| % frontline workers[c] | 73.2 | N/A | 57.3 | 62.6 | 82.2 |
| Number of households | 821 | 27 | 9,215 | 241 | 105 |
| Number of persons | 2,098 | 113 | 18,739 | 619 | 290 |

Behavioral Risk Factor Surveillance System[d]

| Conditions | Native Americans (single race) | | Whites | Latinos | Blacks |
|---|---|---|---|---|---|
| % smokers | 45.5 | - | 20.9 | 21.1 | 33.0 |
| % asthma | 15.2 | - | 11.6 | 12.1 | 8.0 |
| % COPD | 4.8 | - | 2.9 | 4.2 | 2.4 |
| % kidney disease | 3.2 | - | 1.4 | 1.9 | 1.9 |
| % cancer (excl. skin) | 3.4 | - | 3.2 | 3.5 | 1.0 |
| % diabetes | 15.9 | - | 5.2 | 13.7 | 7.9 |
| % heart disease | 3.3 | - | 1.4 | 2.3 | 2.3 |
| % obese (BMI >=30) | 42.5 | - | 28.6 | 34.2 | 30.7 |
| Mean # health cond.[e] | 0.9 | - | 0.5 | 0.7 | 0.5 |
| Min number of persons | 4,708 | - | 24,860 | 587 | 200 |
| Max number of persons | 5,187 | - | 26,720 | 649 | 225 |

#### TEXAS

American Community Survey (ACS)[a]

| | Native Americans (single, homeland) | Native Americans (single, not homeland) | Whites | Latinos | Blacks |
|---|---|---|---|---|---|
| % 3+ generation HH[b] | 2.5 | 4.6 | 2.9 | 7.5 | 4.9 |
| % crowded HH[b] | 2.2 | 3.1 | 2.3 | 11.7 | 4.5 |
| % health ins. (exc. IHS) | 66.5 | 74.4 | 85.4 | 61.6 | 77.4 |
| % health ins. (inc. IHS) | 83.2 | 81.8 | 85.4 | 61.6 | 77.4 |
| Income-poverty ratio | 2.9 | 3.1 | 3.6 | 2.6 | 2.8 |
| % frontline workers[c] | N/A | 60.1 | 49.1 | 71.1 | 62.3 |
| Number of households | 130 | 921 | 168,087 | 101,209 | 32,039 |
| Number of persons | 275 | 2,058 | 352,269 | 252,007 | 72,609 |

Behavioral Risk Factor Surveillance System[d]

| Conditions | Native Americans (single race) | | Whites | Latinos | Blacks |
|---|---|---|---|---|---|
| % smokers | 32.2 | - | 20.4 | 14.6 | 18.5 |
| % asthma | 27.9 | - | 15.0 | 8.7 | 16.9 |
| % COPD | 8.0 | - | 4.2 | 2.5 | 4.5 |
| % kidney disease | 4.8 | - | 1.8 | 2.3 | 2.3 |
| % cancer (excl. skin) | 6.9 | - | 3.9 | 2.2 | 2.5 |
| % diabetes | 11.2 | - | 6.1 | 11.2 | 9.8 |
| % heart disease | 2.6 | - | 1.6 | 1.4 | 2.2 |
| % obese (BMI >=30) | 35.4 | - | 28.6 | 37.4 | 38.6 |
| Mean # health cond.[e] | 1.0 | - | 0.6 | 0.7 | 0.8 |
| Min number of persons | 442 | - | 27,980 | 17,396 | 4,672 |
| Max number of persons | 481 | - | 30,189 | 20,340 | 5,234 |

WASHINGTON

American Community Survey (ACS)[a]

| | Native Americans (single, homeland) | Native Americans (single, not homeland) | Whites | Latinos | Blacks |
|---|---|---|---|---|---|
| % 3+ generation HH[b] | 6.9 | 4.2 | 2.7 | 5.7 | 3.8 |
| % crowded HH[b] | 8.5 | 6.6 | 2.6 | 14.4 | 7.7 |
| % health ins. (exc. IHS) | 80.0 | 85.5 | 93.6 | 72.6 | 88.9 |
| % health ins. (inc. IHS) | 96.6 | 92.5 | 93.6 | 72.7 | 88.9 |
| Income-poverty ratio | 2.5 | 2.6 | 3.6 | 2.6 | 2.9 |
| % frontline workers[c] | 65.2 | 55.6 | 51.2 | 74.6 | 61.4 |
| Number of households | 1,365 | 260 | 68,150 | 8,028 | 2,449 |
| Number of persons | 3,210 | 674 | 145,501 | 20,642 | 6,076 |

Behavioral Risk Factor Surveillance System[d]

| Conditions | Native Americans (single race) | | Whites | Latinos | Blacks |
|---|---|---|---|---|---|
| % smokers | 37.9 | - | 18.0 | 12.3 | 18.1 |
| % asthma | 23.4 | - | 17.5 | 12.0 | 15.6 |
| % COPD | 7.5 | - | 3.9 | 2.7 | 2.5 |
| % kidney disease | 2.8 | - | 1.8 | 2.8 | 2.6 |
| % cancer (excl. skin) | 6.3 | - | 3.9 | 2.0 | 2.1 |
| % diabetes | 9.2 | - | 5.8 | 11.5 | 9.7 |
| % heart disease | 2.7 | - | 1.2 | 1.4 | 1.6 |
| % obese (BMI >=30) | 34.0 | - | 27.6 | 34.5 | 33.7 |
| Mean # health cond.[e] | 0.8 | - | 0.6 | 0.7 | 0.7 |
| Min number of persons | 772 | - | 43,815 | 4,944 | 1,299 |
| Max number of persons | 877 | - | 47,466 | 6,219 | 1,455 |

#### WISCONSIN

American Community Survey (ACS)[a]

| | Native Americans (single, homeland) | Native Americans (single, not homeland) | Whites | Latinos | Blacks |
|---|---|---|---|---|---|
| % 3+ generation HH[b] | 3.5 | 2.1 | 1.5 | 4.3 | 3.7 |
| % crowded HH[b] | 3.7 | 3.1 | 1.5 | 8.9 | 4.6 |
| % health ins. (exc. IHS) | 75.2 | 81.7 | 94.1 | 73.9 | 85.5 |
| % health ins. (inc. IHS) | 94.2 | 86.6 | 94.1 | 74.0 | 85.5 |
| Income-poverty ratio | 2.4 | 3.1 | 3.5 | 2.5 | 2.2 |
| % frontline workers[c] | 67.7 | N/A | 57.6 | 74.0 | 70.4 |
| Number of households | 519 | 88 | 65,089 | 2,568 | 2,341 |
| Number of persons | 1,203 | 262 | 136,774 | 6,519 | 5,886 |

Behavioral Risk Factor Surveillance System[d]

| Conditions | Native Americans (single race) | | Whites | Latinos | Blacks |
|---|---|---|---|---|---|
| % smokers | 36.8 | - | 20.4 | 16.9 | 30.5 |
| % asthma | 16.3 | - | 13.1 | 12.0 | 22.1 |
| % COPD | 3.5 | - | 3.1 | 3.9 | 5.2 |
| % kidney disease | 2.6 | - | 1.2 | 1.6 | 2.6 |
| % cancer (excl. skin) | 4.7 | - | 3.1 | 2.6 | 1.9 |
| % diabetes | 9.2 | - | 4.7 | 9.7 | 11.6 |
| % heart disease | 2.4 | - | 1.3 | 0.8 | 2.3 |
| % obese (BMI >=30) | 40.1 | - | 29.0 | 35.0 | 39.9 |
| Mean # health cond.[e] | 0.8 | - | 0.6 | 0.7 | 0.9 |
| Min number of persons | 586 | - | 21,696 | 899 | 1,516 |
| Max number of persons | 639 | - | 23,422 | 1,064 | 1,672 |

*Note: N/A indicates number of households or number of persons is below 50.*

[a] Based on respondents ages 18-60 (except for estimates of frontline workers).
[b] This estimate is based on information at the household-level; the remaining ACS

variables are based on individual information.
[c] Based on persons of all ages reporting jobs in the past five years from the single-year 2018 ACS.
[d] Based on respondents ages 18-59. The number of persons providing responses in the BRFSS varies across health conditions. The minimum sample size pertains to persons with responses to all of the health condition variables.
[e] The mean number of health conditions per person is the sum of affirmative responses to the following conditions: asthma, COPD, kidney disease, cancer, diabetes, heart disease, and obesity (BMI≥30).

**Appendix Table 2: Correlations between selected risk factors for COVID-19 infection and mortality and the Standardized Mortality Ratio among Native Americans across states**

| | SMR (COVID-19, 2020)[a] | SMR (all cause, 2019)[a] |
|---|---|---|
| % 3+ generation HH | 0.77 | 0.66 |
| % crowded HH | 0.60 | 0.63 |
| % health insurance (exc. IHS) | -0.79 | -0.78 |
| % health insurance (inc. IHS) | 0.48 | 0.47 |
| Income-poverty ratio | -0.84 | -0.77 |
| % frontline workers | 0.54 | 0.52 |
| % smokers | 0.19 | 0.46 |
| % asthma | -0.52 | -0.49 |
| % COPD | -0.50 | -0.34 |
| % kidney disease | -0.01 | -0.12 |
| % cancer (excl. skin) | -0.49 | -0.48 |
| % heart disease | -0.21 | 0.02 |
| % diabetes | 0.65 | 0.39 |
| % obese (BMI >= 30) | 0.55 | 0.39 |
| Average openness score | 0.41 | 0.57 |

[a] SMRs are based on indirect standardization by age for Native Americans. The SMR for COVID-19 deaths in 2020 uses the same data as in the main analysis. The SMR for all-cause mortality in 2019 uses CDC's estimates of all-cause mortality in 2019 (https://wonder.cdc.gov/).

**Appendix Figure 1: Average (mean) age by race and ethnicity for 16 states**

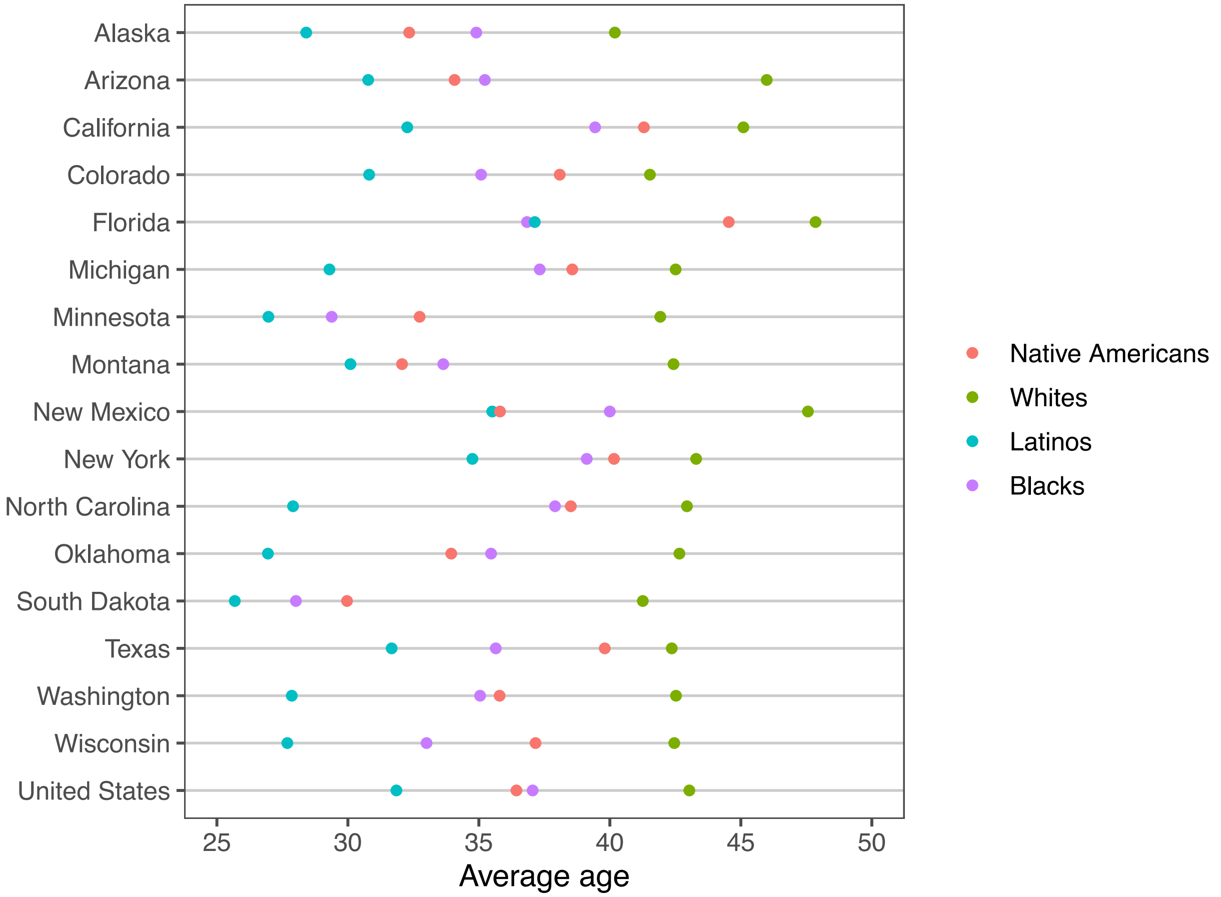


Source: ACS 5-year estimates, 2015-2019

**Appendix Figure 2: Standardized Mortality Ratios for COVID-19 and 95% confidence intervals by race and ethnicity for 16 states**

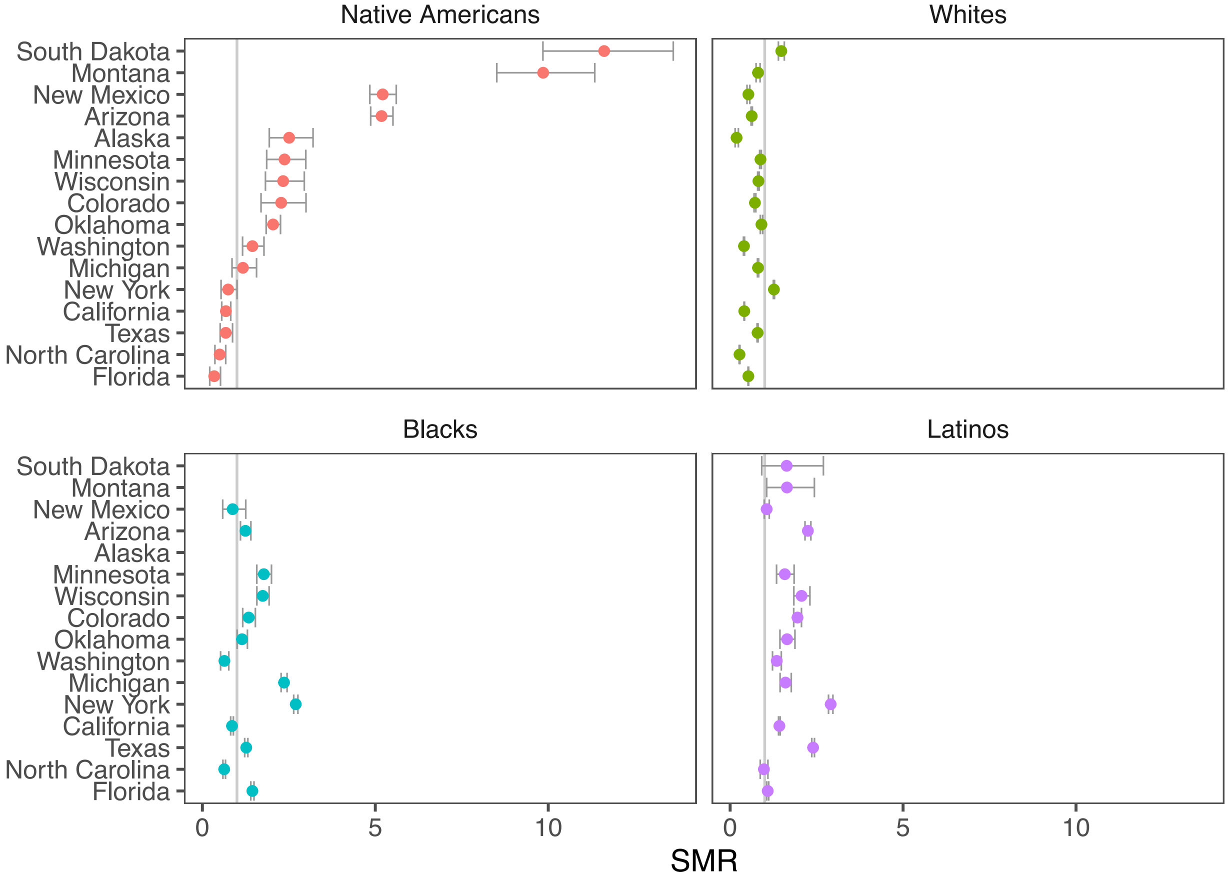


Source: CDC Weekly Updates by Select Demographic and Geographic Characteristics (as of Jan. 21, 2021)

*Note: SMRs are based on indirect standardization by age; 95% confidence intervals (95%) are calculated according to the procedure presented in Ulm (1990).*
